# Replay of Interictal Sequential Activity Shapes the Epileptic Network Dynamics

**DOI:** 10.1101/2024.03.28.24304879

**Authors:** Kang Wang, Haixiang Wang, Yuxiang Yan, Wenzheng Li, Fang Cai, Wenjing Zhou, Bo Hong

## Abstract

Both the imbalance of neuronal excitation and inhibition, and the network disorganization may lead to hyperactivity in epilepsy. However, the insufficiency of seizure data poses the challenge of elucidating the network mechanisms behind the frequent and recurrent abnormal discharges. Our study of two extensive intracranial EEG datasets revealed that the seizure onset zone exhibits recurrent synchronous activation of interictal events. These synchronized discharges formed repetitive sequential patterns, indicative of a stable and intricate network structure within the seizure onset zone (SOZ). We hypothesized that the frequent replay of interictal sequential activity shapes the structure of the epileptic network, which in turn supports the occurrence of these discharges. The Hopfield-Kuramoto oscillator network model was employed to characterize the formation and evolution of the epileptic network, encoding the interictal sequential patterns into the network structure using the Hebbian rule. This model successfully replicated patient-specific interictal sequential activity. Dynamic change of the network connections was further introduced to build an adaptive Kuramoto model to simulate the interictal to ictal transition. The Kuramoto oscillator network with adaptive connections (KONWAC) model we proposed essentially combines two scales of Hebbian plasticity, shaping both the stereotyped propagation and the ictal transition in epileptic networks through the interplay of regularity and uncertainty in interictal discharges.

## Introduction

Epilepsy affects nearly 65 million people worldwide, with one-third of patients unable to effectively control their seizures with medication. The high costs and mortality associated with epilepsy underscore the urgent need for a deeper understanding of the nature of epilepsy^1^. In recent years, epilepsy has garnered growing attention as a network disorder rather than a localized anomaly^2,3^. This network perspective has led to numerous new theories and approaches for the diagnosis and treatment of epilepsy, particularly in identifying the critical nodes or network topological properties of epileptic networks^4–12^. In addition, a growing body of research has begun to unravel the dynamic interaction between nodes within epileptic networks and how their interactions significantly influence the manifestation of epileptic seizures^4,13–18^. Identifying key nodes within epileptic networks and analyzing their interactions will help neurosurgeons maximize the benefits of surgical or neurostimulation interventions while minimizing associated harm^19–21^. Prior research has primarily focused on characterizing the structure of epileptic networks^22–24^ and conducting statistical analyses of network node activity patterns. However, there has been a lack of investigation into the network dynamics underlying epileptic activity^12,25,26^.

Revealing the dynamics within epileptic networks relies on the availability of sufficient neurophysiological data to mitigate the interference of noise in mechanism exploration. Seizures, being the most prominent pathological activity in epilepsy, are often used as standard diagnostic data in clinical settings^27^. However, it is costly to obtain an adequate amount of seizure data due to limited clinical observation time and risk of infection in the case of intracranial recording of seizure EEG. This problem becomes particularly pronounced when there is a need for personalized modeling of patients’ brain networks. With the widespread use of intracranial EEG (iEEG) in the evaluation of epilepsy, substantial interictal data can often be obtained, providing valuable pathological information^28–31^. Interictal events (IEs), including high frequency oscillations (HFOs) and spikes, are considered important biomarkers in epilepsy. Their value in addressing issues such as seizure onset zone (SOZ) localization has been validated^32–36^. Furthermore, the approximate synchrony of recurrent interictal electrographic events across different nodes provides rich spatiotemporal information about the epileptic network^22,37–39^. The earlier discharges within these collective activities have a higher probability to be in the epileptic origin^23,40,41^. These recurrent interictal activities not only serve as representative samples of epileptic network activity ^42^, but are also speculated to potentially drive the formation and evolution of the epileptic network^12^. Moreover, the long-term periodic coupling of their occurrence rates with seizures underscores their pivotal role in parsing the epilepsy network, paralleling seizure activity^43,44^. This study, therefore, aims to harness the wealth of information inherent in interictal synchronized population activity, to deepen our understanding of the intricacies of epilepsy networks. We aim to reveal the core structure of these networks and elucidate the rich dynamics that culminate in epileptic seizures. In this study, we recorded continuous stereoelectroencephalography (SEEG) data for an average of 24 hours per patient from 18 individuals with drug-resistant epilepsy, for a total of 448 hours of interictal data and 77 seizures. To validate the generalizability of our epilepsy network analysis methods, we also conducted a parallel analysis using the Epilepsiae dataset^45^. This dataset comprises continuous intracranial EEG recordings of over 100 hours on average per patient from 20 individuals, including a total of 3683.9 hours of interictal data and 399 seizures. The IEs extracted from both datasets prove effective in accurately identifying the SOZ in individual patients. Moreover, the IEs within the SOZ exhibit a consistent and repetitive propagation pattern, indicating the presence of intricate network structures within the SOZ. Our analysis of the temporal characteristics of interictal event revealed a clear periodicity. This periodic and stereotyped propagation of interictal events beared resemblance to the phenomenon of “memory replay” in functional brain networks and inspireed our hypothesis about the network mechanisms underlying the formation and maintenance of interictal repetitive activity. We employed the Kuramoto oscillator model to simulate the stereotyped repetitive interictal population activity, and adopted the Hebbian rule to determine the strength of connections between oscillator nodes^46^. This approach successfully replicated individualized interictal population activities. Furthermore, in order to elucidate the mechanisms by which interictal activity evolves into seizure events, we treated interictal and ictal states as two distinct states of a dynamical system and conducted a joint network dynamics analysis. We discovered that the dynamic plasticity of network connections, as demonstrated in the oscillator model^47^, can lead to a stochastic phase transition from interictal activity to epileptic seizures. The Kuramoto oscillator network with adaptive connections (KONWAC), proposed in this study, offers a network dynamics perspective that elucidates the regularity of stereotyped repetition in epileptic networks and the uncertainty in seizure evolution. This model provides a novel framework for diagnosis and intervention based on epileptic network dynamics.

## Results

### 1. Coactivated Interictal Events within the SOZ Form Reproducible Sequence

Drawing on the high-frequency features of HFOs (mainly 80-250Hz) and the broadband characteristics of spikes, we utilized the high-frequency energy within the 80-250Hz range as a unified feature for detecting interictal events. As shown in Fig. 1, the distribution of detected interictal events closely correlates with the electrodes in the clinically identified seizure onset zone (SOZ), suggesting that “high-frequency energy interictal events” may serve as a biomarker of epileptic network activity (average AUC of 0.857 for Yuquan and average AUC of 0.772 for Epilepsiae; Fig. 1b). Adopting the approach of previous studies^48^, we manually annotated three types of interictal events detected in patient Y1 (HFOs, spikes, and a coupled type of HFO riding on spike), which appeared to be representative of interictal events (Supplementary Fig. 1). Each of these events exhibited a specific enhancement of high-frequency energy (>50Hz) (Supplementary Fig. 2). This observation supports the use of high-frequency energy interictal events as a basis for investigating the dynamics of epileptic networks, temporarily disregarding morphological differences among interictal events.

**Fig. 1.**
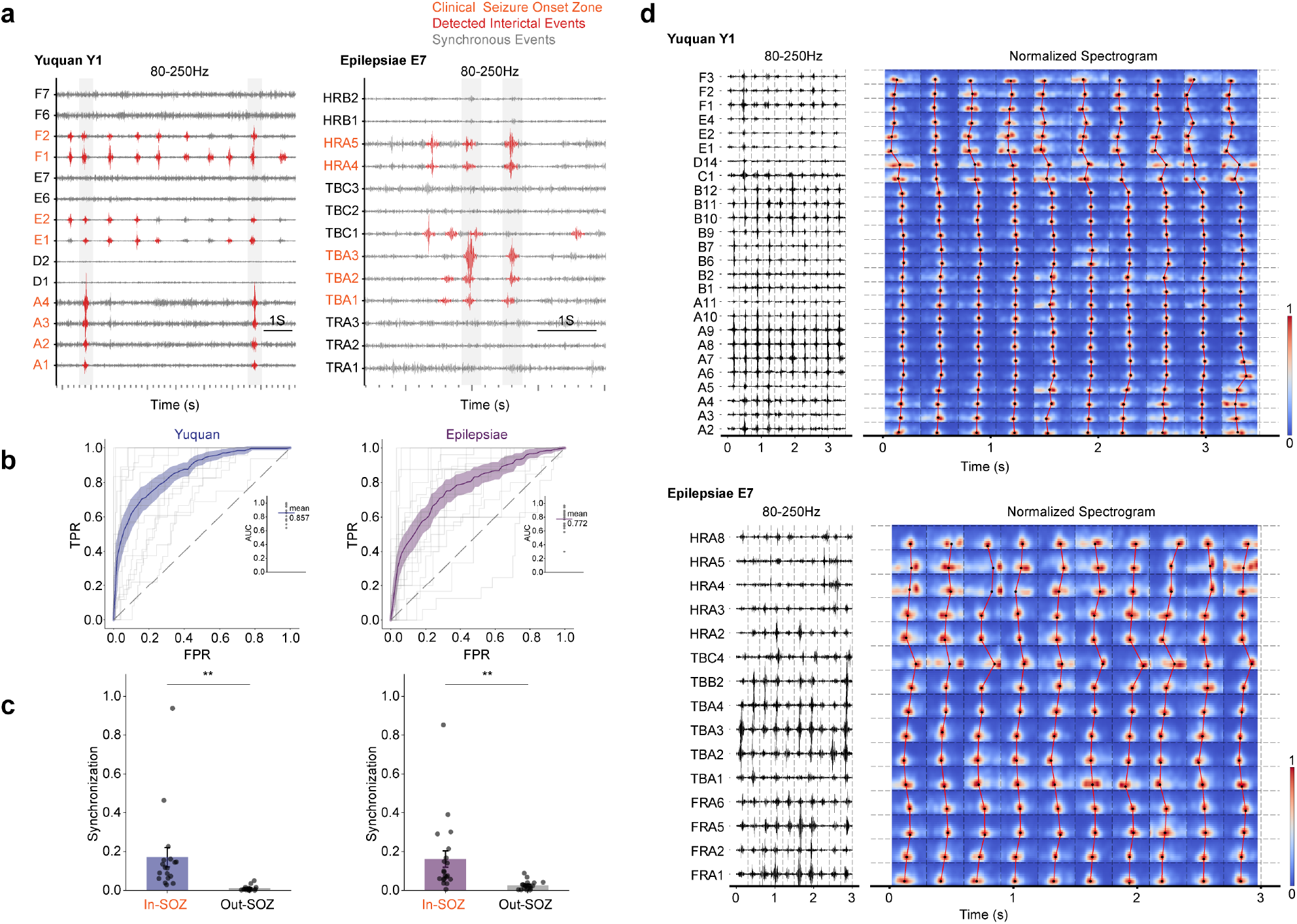
Coactivated interictal events within the SOZ with reproducible propagation. **a**, Detected interictal events and population events. The interictal SEEG signal filtered with an 80-250Hz band-pass filter is shown with clinical SOZ, detected interictal events, and population events annotated. **b**, Agreement between clinical SOZ and SOZ prediction based on interictal event distribution. The ROC curve was used to evaluate the prediction performance in both the Yuquan and Epilepsiae datasets. The mean ROC curve is highlighted in color, while the shaded region indicates the s.e.m. In addition, individual patient ROC curves are represented by bright lines in the background. Patient AUC values are shown in the insets, with the mean AUC indicated by a horizontal line. **c**, Coactivation level of interictal events within and outside the clinical SOZ. The degree of synchronization reflects the extent to which interictal events overlap between any two contacts within a region. Mean and s.e.m. are presented as bars and error bars, respectively. Wilcoxon signed-rank test was performed to compare the degree of synchronization within and outside the clinical SOZ (**p<0.01). **d**, Synchronously activated interictal events exhibit repetitive propagation patterns. Ten representative population activities from two patients Y1 and E7 are presented, featuring filtered 80-250Hz high-frequency activity and corresponding normalized time-frequency maps, where the black dots indicate the energy centroids of the maps. These patterns suggest a consistent mode of propagation in interictal events.

**Fig. 2.**
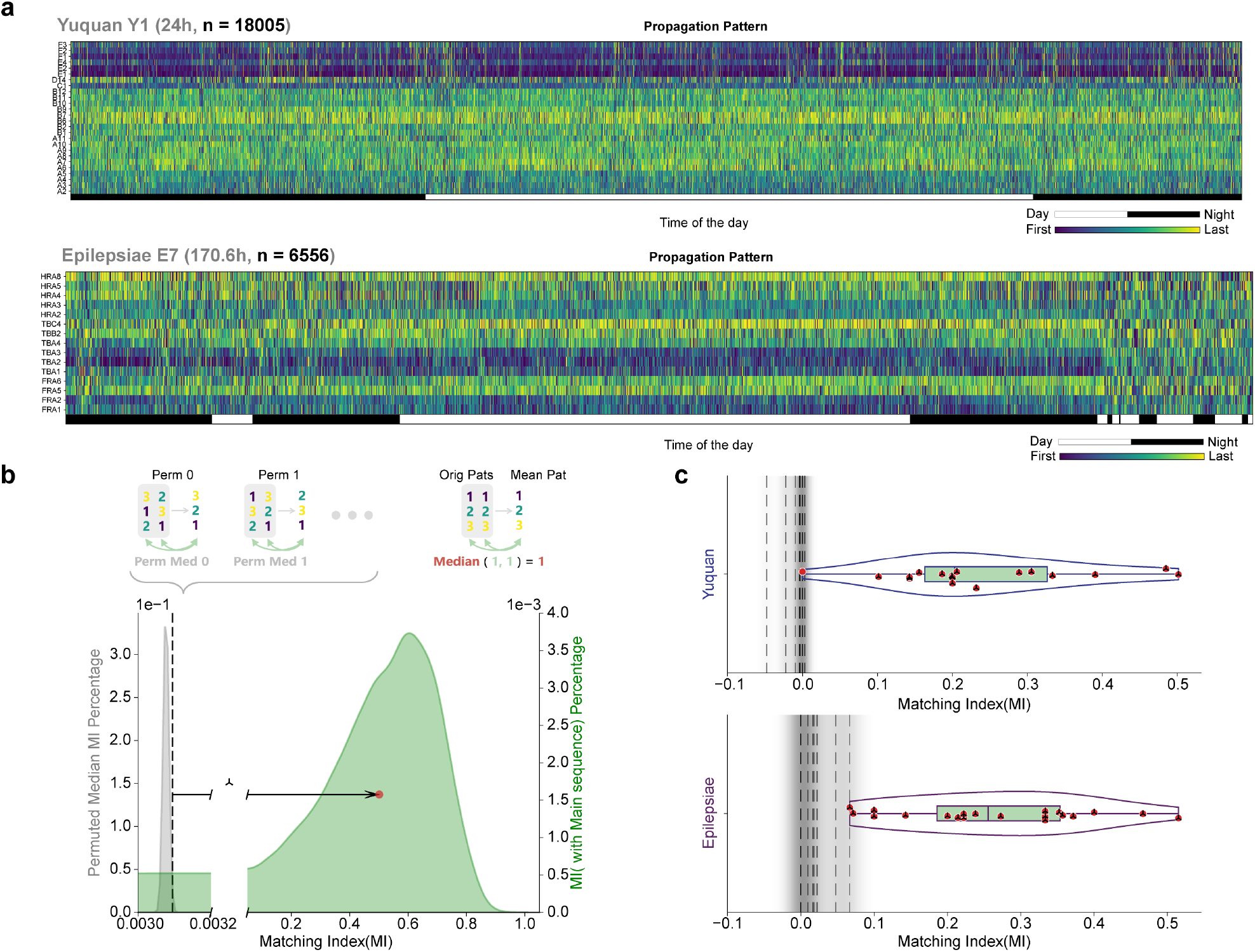
Stable reproducibility of interictal propagation across patients. **a**, Stable and stereotyped propagation patterns in long-term data of Yuquan patient Y1 (24h continuous SEEG, population events n=18005) and Epilepsiae patient E7 (6556 population events over 170.6 hours of iEEG data). Each column represents one propagation pattern. The average propagation pattern projected onto contacts can be found in Supplementary Fig. 3. **b**, A permutation method was used to test the statistical significance of the propagation reproducibility. The true MI distribution (green area) was obtained by comparing all propagation patterns with their average pattern. Reproducibility is considered significant if the real MI median (red dots) lies to the right of the significance threshold (black dotted line, 95% cumulative probability, p<0.05) of a surrogate distribution of the MI median (gray area, propagation patterns permutated 200 times). **c**, The results of the reproducibility test for all patients in the Yuquan and Epilesiae datasets. All patients in the Yuquan and Epilesiae datasets, except for one patient in the Yuquan dataset, show significant reproducibility of interictal propagation patterns, with the red dots and black dotted lines having the same meaning as in **b**.

These interictal events were synchronously activated across multiple SOZ contacts (Fig. 1a). We compared the degree of synchrony (the temporal overlap of interictal events between pairs of contacts) within and outside the SOZ, and found significantly stronger synchrony within the SOZ (as determined by the Wilcoxon signed-rank test, p<0.01 for both the Yuquan and Epilepsiae datasets; Fig. 1c). Incorporating this synchrony constraint of abnormal discharges into the detection of interictal events also significantly improved the prediction of SOZ in the Yuquan dataset (mean AUC 0.89 under synchrony constraints, p=0.023, based on paired t-test; see Supplementary Fig. 3a,b). Overall, this indicates a strong link between the SOZ and interictal populational activity. Within these interictal activities, we further observed the propagation of interictal events between different sites, exhibiting a reproducible sequence (Fig. 1d).

**Fig. 3.**
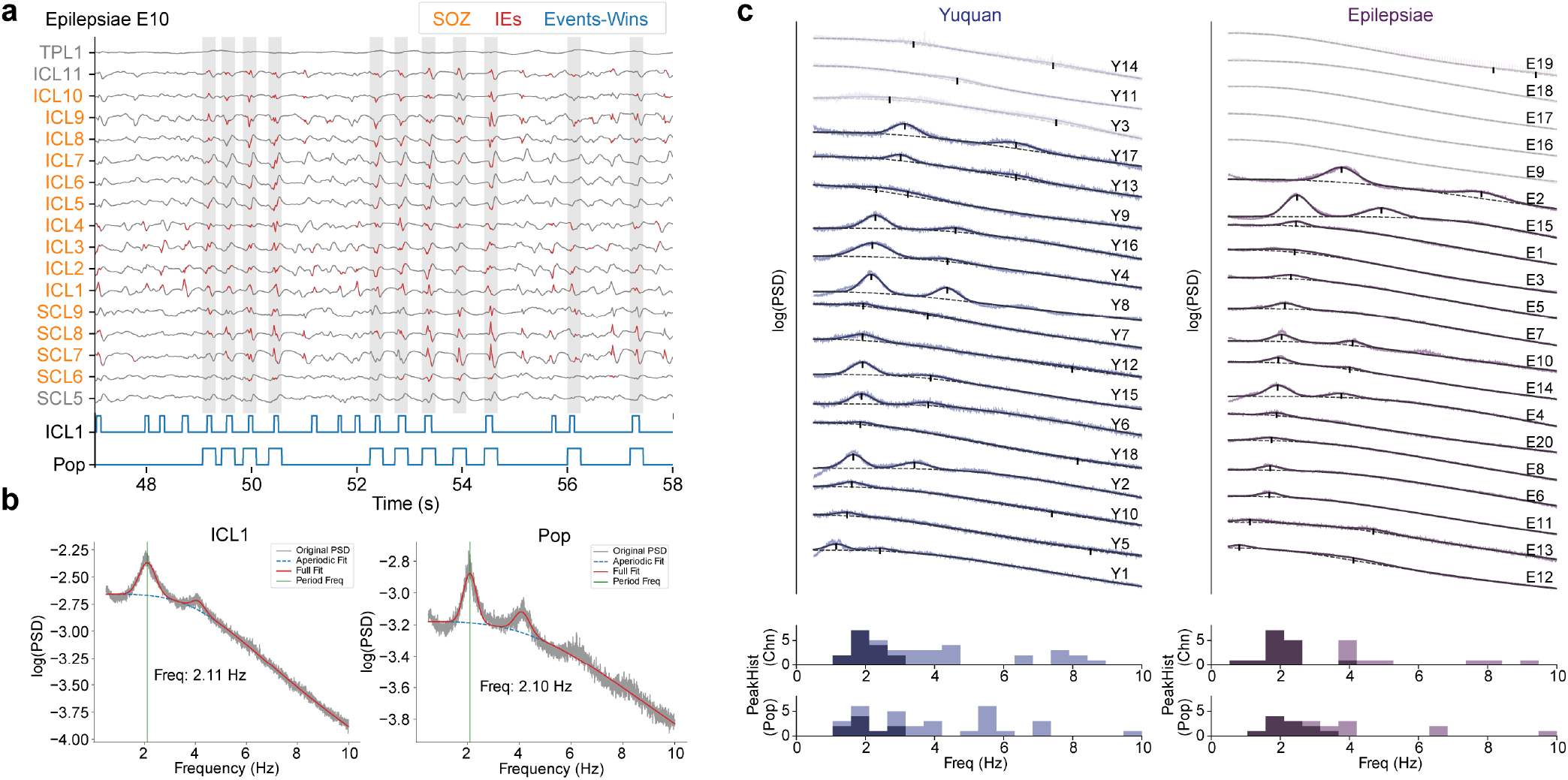
Periodicity of local and population interictal events. **a**, Example data illustrating the periodicity of interictal events, where the orange represents the clinically annotated SOZ contacts, red and gray represent the detected local and population interictal events, respectively. The signals of the occurrence time windows for population events or local events at the ICL1 contact are shown below, with event occurrences marked by elevated signals. **b**, The spectrum of the occurrence time window signals for interictal events at the ICL1 contact and population-level. Fitting and separating the spectrum into periodic and non-periodic components reveals a distinct periodic component around 2Hz. **c**, Periodicity of interictal events in patient data. For each dataset, the spectral decomposition results of event occurrence signals from representative electrodes of each patient are arranged and presented, with each part corresponding to those in **b**. The frequencies of the periodic components of the representative electrodes are displayed in the histogram below, with significant periodic components within 0.5-4Hz being highlighted in the histogram and in the above spectrums with dark color. The periodicity of the population activities is also shown below. It can be observed that the periodic frequency of the interictal events is around 2Hz.

The recurrent manifestation of interictal population activities may arise from the stable network structure within the SOZ, which in turn further indicates that the recurrent interictal activities might exhibit certain regularities. For each population event, we characterized the propagation pattern using the time-frequency centroids of the 80-250Hz bandpass signals of the participating contacts (see Methods). This approach enabled the observation of stable and repetitive propagation patterns across numerous interictal population events. In the 24-hour SEEG data of patient Y1 from the Yuquan dataset, we identified 18,005 population events, whose propagation patterns were repetitive and maintained long-term stability throughout the whole day (Fig. 2a). By integrating electrode locations, the stable interictal propagation patterns also revealed fine spatial organization within the SOZ. For example, in patient Y1, a propagation trend from the insular cortex to the frontal lobe and cingulate gyrus was observed (Supplementary Fig. 4). Similarly, in patient E7 from the Epilepsiae dataset, we extracted 6,556 population events from 170.6 hours of continuous iEEG data, which also showed a repetitive propagation sequence from the anterior to the posterior temporal region (Fig. 2a; Supplementary Fig. 4).

**Fig. 4.**
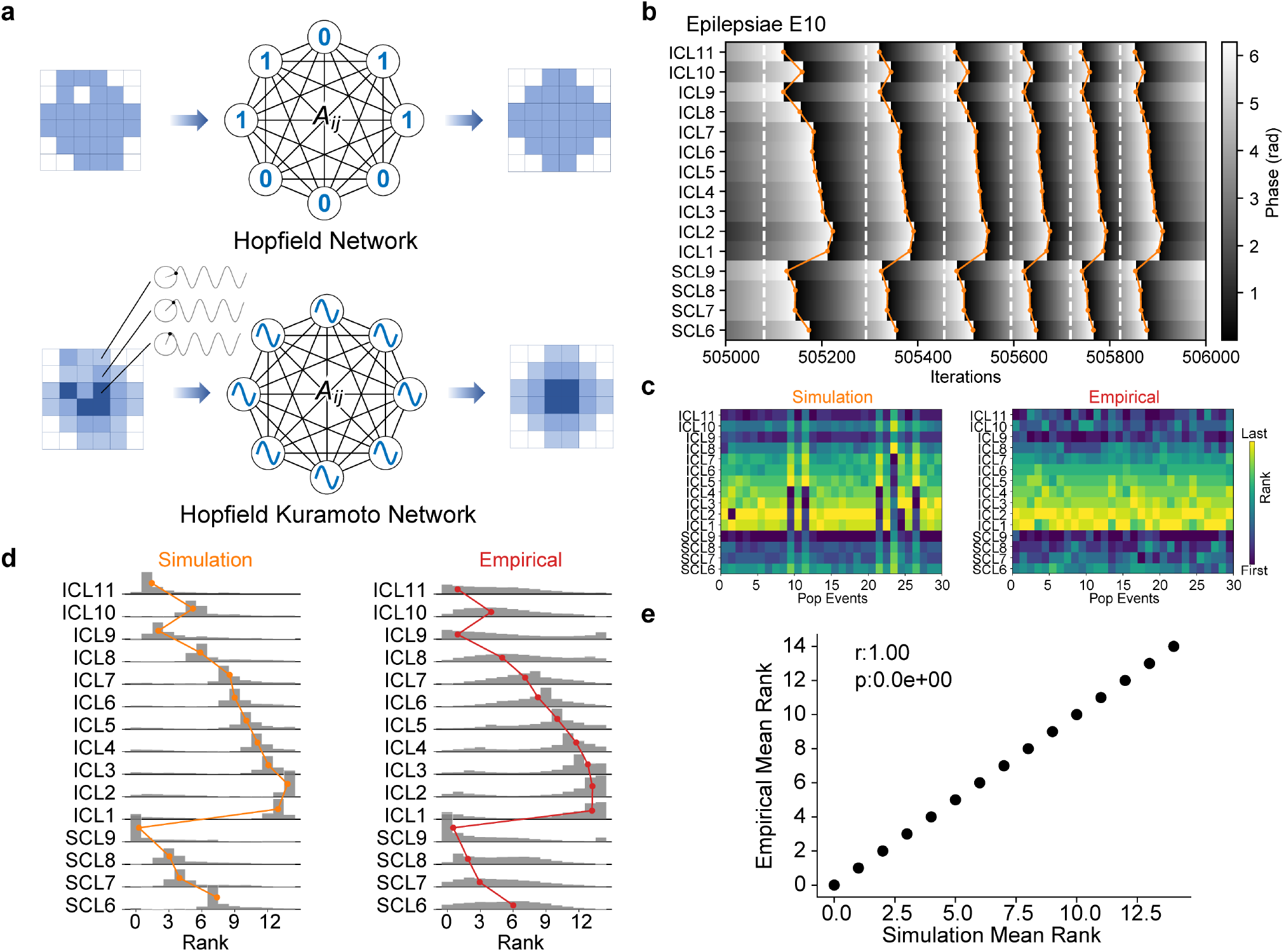
The Hopfield Kuramoto model encodes specific phase patterns. **a**, The traditional Hopfield model can encode specific signal patterns, while the Hopfield Kuramoto model can encode specific phase order patterns. **b**, Based on the concept of the Hopfield network and using a method similar to Hebbian rule to construct a network encoding specific phase patterns, the model can generate patient-specific interictal propagation patterns for patient E10. In the Kuramoto model, we posit that whenever a node phase reaches approximately 2*π*, an interictal event is considered to have occurred. The orange line indicates the sequence of each propagation, and the white dashed line represents the boundary between population activities. **c**, Sample propagation patterns of interictal population activities in simulated or real data. **d**, Statistical distribution of each node’s position in the propagation patterns in simulated or real data. **e**, The Spearman correlation of the average propagation patterns in simulated and real data, indicated by the colored lines in **d**. The Hopfield Kuramoto network is able to effectively encode and explain patient-specific interictal propagation patterns.

To quantify and test the significance of the aforementioned sequence repetitiveness, we assumed that a more repetitive propagation pattern will yield a clearer and more consistent average sequence. For an individual patient, we can assess the overall repetitiveness of their propagation sequences by calculating the consistency between their average propagation sequence and all individual propagation events, and then extend this to a group level. We used the Matching Index (MI)^22^ to compute the consistency between two propagation sequences, mapping opposite or consistent pairs of propagation sequences onto a -1 to 1 scale (see Methods). The better the repeatability of a patient’s propagation sequences, the more the distribution of MI values between individual and average sequence will be right-skewed towards 1, with the median of this distribution quantifying the quality of propagation repetitiveness. To test whether the repeatability of the patient’s propagation patterns is significant, we checked whether this MI median lies in the rightmost 5% of its surrogate distribution (permutation test) (Fig. 2b). The results indicated significant repetitiveness of interictal propagation sequence at the group level (with a significant proportion of 17/18 for Yuquan patients and 20/20 for Epilepsiae patients; Fig. 2c).

We further examined the temporal, frequency, and spatial characteristics of these repetitive propagation activities. Firstly, we calculated the delay between temporally adjacent sites during propagation, showing that propagation between electrodes is remarkably rapid, approximately around 2ms, akin to the timescale of a synapse (the overall median propagation delay for the Yuquan dataset was 3.37 [1.28-8.01]ms, and 4.50 [1.66-11.04]ms for the Epilepsiae dataset, median [25th quantile-75th quantile]; Supplementary Fig. 5). Despite the fast propagation, the characteristics of stable propagation patterns were still discernible, demonstrating the effectiveness of our method based on time-frequency centroids. Secondly, we observed that the overall centroid frequency was around 110Hz. However, due to 1/f power-law spectral characteristics in some broadband activities, this centroid frequency might be lower than actual (the overall median of centroid frequencies for the Yuquan dataset was 107.69 [101.33-115.36] Hz, and for the Epilepsiae dataset was 109.20 [102.04-119.04] Hz; Supplementary Fig. 5). Lastly, contrary to the traditional view of the SOZ as a homogeneous region, stable interictal propagation sequence revealed more detailed distinctions between electrodes or regions within the SOZ. As shown in Supplementary Fig. 6a, for two patients whose lesions involved the insula, frontal lobe, and cingulate gyrus, similar propagation paths were observed, with propagation from the insula to the frontal lobe and later involving the cingulate gyrus. The distribution of upstream electrodes (the first 50% in propagation) across different brain areas also suggests a common insular origin for the SOZ in both patients (Supplementary Fig. 6a). Our spatial analysis of interictal propagation patterns at the group level is consistent with the etiology of the patients. The results indicate that interictal propagation in Yuquan patients mostly originate from the frontal, parietal, and insular lobes, whereas in Epilepsiae patients, often originate from the temporal lobe, limbic system, and frontal lobe (Supplementary Fig. 6b). This observation aligns with the fact that most Yuquan patients have focal cortical dysplasia (FCD) and tuberous sclerosis complex (TSC) etiologies, and most Epilepsiae patients have temporal lobe epilepsy, such as hippocampal sclerosis. These findings suggest that stable interictal propagation sequence can provide a detailed depiction of the SOZ, helping to identify more critical nodes or origin sites within it.

**Fig. 5.**
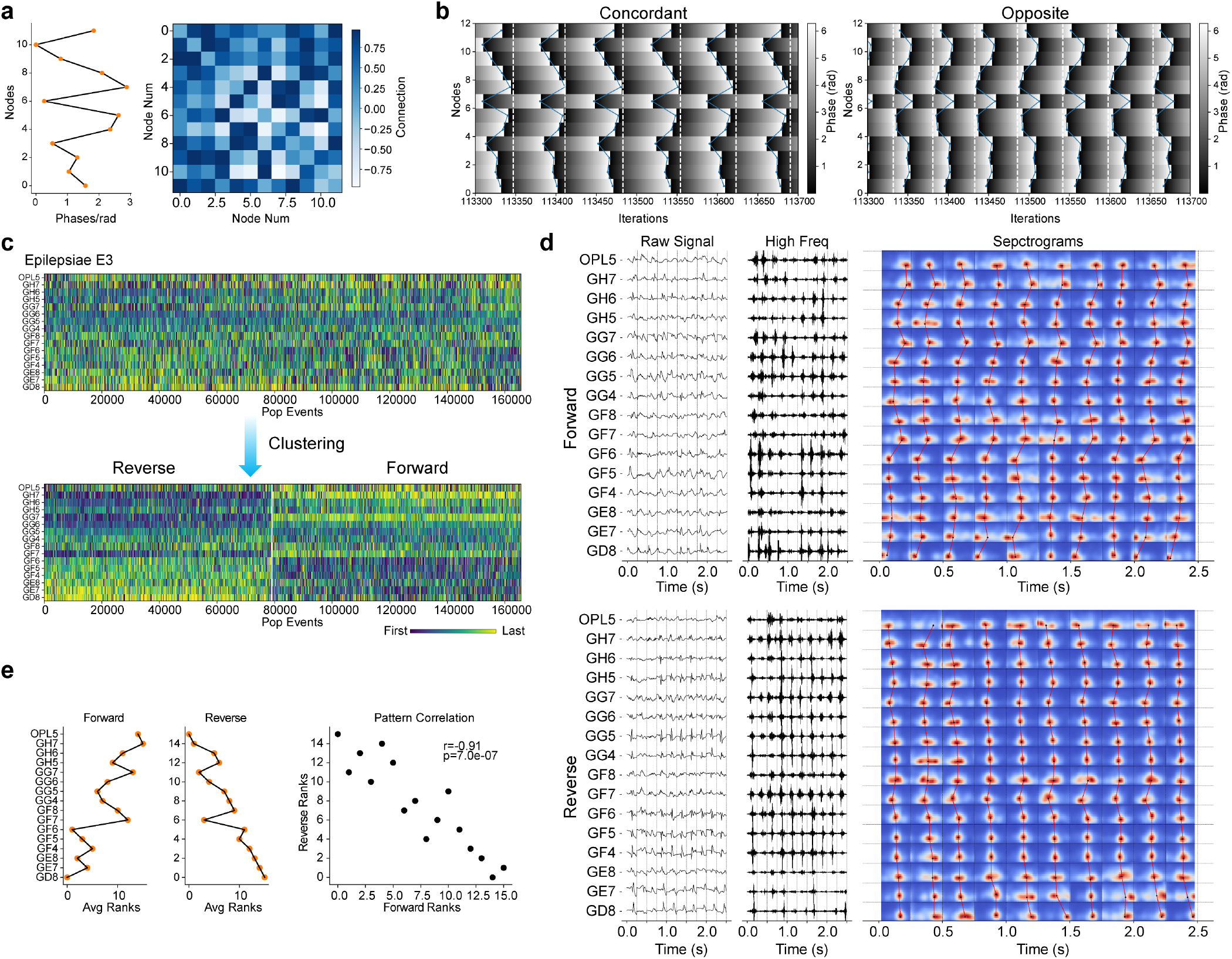
Symmetric network structures based on Hebbian rule can simultaneously encode opposite propagation patterns. **a-b**, Simulation of Hebbian network simultaneously encoding forward and reverse propagation patterns. **a**, The preset propagation pattern and Hebbian network structure derived from the preset pattern. **b**, For the same network structure in **a**, network activity can converge to propagation patterns that are either concordant with or opposite to the preset pattern. The two convergent propagation patterns are shown, where white dotted lines delineate the boundaries of population activities and blue lines represent the propagation patterns. **c-e**, Patient E3’s data show two interictal propagation patterns with opposite directions, supporting the phase order encoding model of the symmetric Hebbian network structure. **c**, Samples of interictal propagation patterns of patient E3, where clustering produces two classes of clearer propagation patterns. **d**, The typical population activities corresponding to the two classes of propagation patterns in **c. e**, There is a significant negative Spearman correlation between the average propagation patterns of the two types of propagation activities in **c**.

**Fig. 6.**
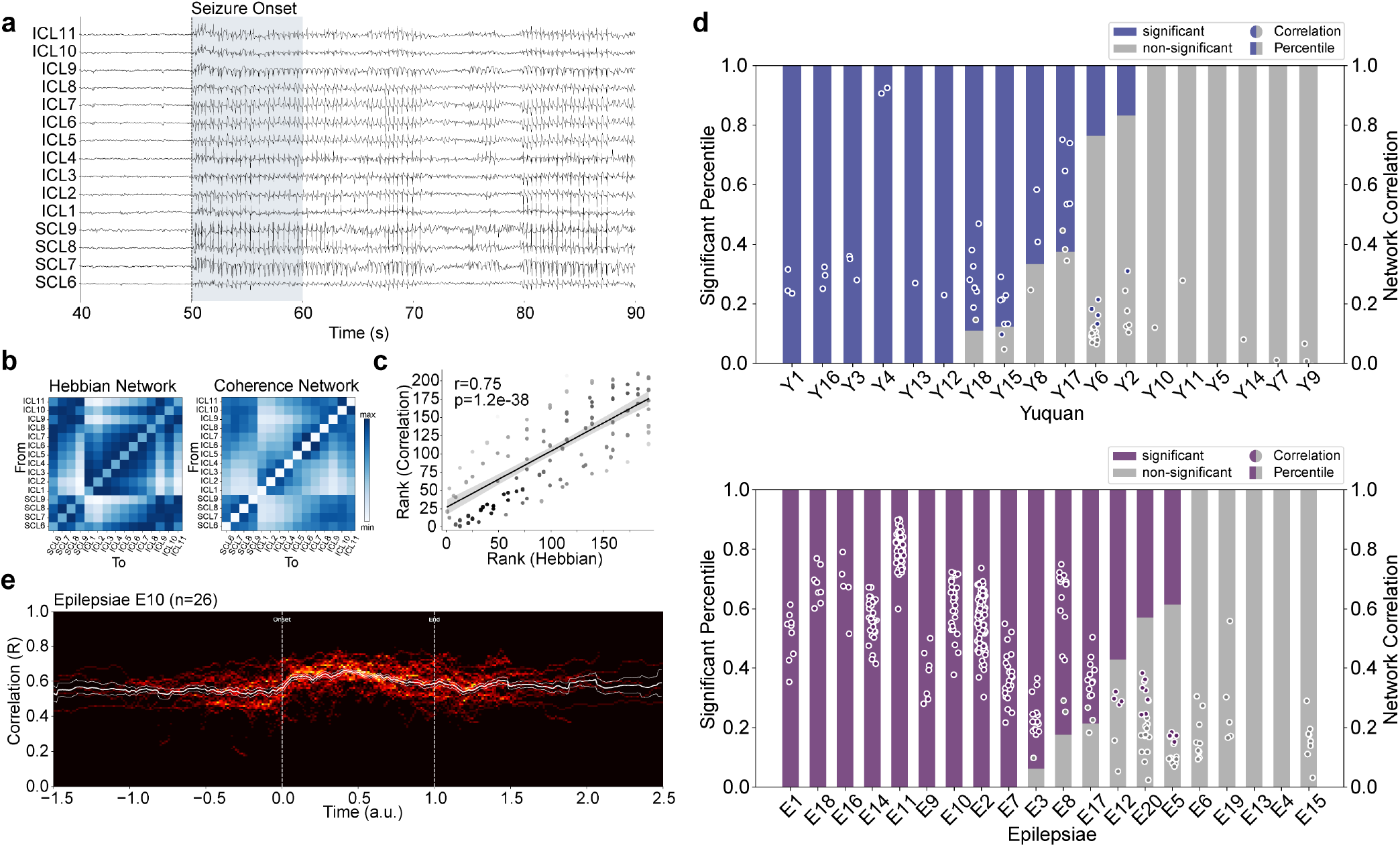
Interictal Hebb network carries seizure initiation. **a**, Raw ictal signal recorded from contacts involved in the interictal propagation of Epilepsiae patient E10. **b**, The ictal coherence network and the interictal Hebb network are shown. The former was calculated using the average coherence of 1-250Hz at seizure onset (shaded area in **a**). **c**, Spearman correlation was calculated between the ictal network and the interictal network (panel **b**, excluding diagonal elements) to assess network consistency. The shaded area represents the 95% confidence interval, while the brightness of each point reflects the density of points around it. The correlation value and p-value are shown in the upper left. **d**, Consistency between interictal and ictal networks across patients is shown for the Yuquan dataset (top) and the Epilepsiae dataset (bottom), with colored bars representing the proportion of seizures whose onset networks significantly match the interictal network. Corresponding correlation values for these seizures are shown as colored dots. Results are FDR corrected for each patient, with significant set as p<0.05. Some negative correlation points are not shown. **e**, Continuous changes in network consistency for patient E10. The background shows the superposition of network consistency change curves of multiple seizures (aligned by start and end time) in a two-dimensional histogram, with white lines indicating the mean and s.e.m. Network consistency is particularly enhanced near seizure onset, suggesting the contribution of interictal network to seizure initiation. Subsequent consistency variation indicates some plastic structural changes during seizures.

### 2. Periodicity and Stochasticity of Interictal Events Occurrence

One of the distinguishing characteristics of epileptic activity from normal physiological activity is its strong spontaneity. This spontaneity indicates an underlying self-sustaining, stable structure that supports the cyclical switching between different states. Understanding the regularities in these state transitions may help unravel the dynamic mechanisms in epileptic networks. Thus, we transformed the interictal events into discrete event sequences to study the temporal regularities in the occurrence of interictal events. This was achieved by marking the moments of interictal event occurrences in the signal, thus obtaining time stamps of these events (as shown at the bottom of Fig. 3a). A clear periodicity was shown in the occurrence of interictal events (Fig. 3a). By performing spectral decomposition on the time stamps of population or local interictal discharges, we were able to extract and quantify the periodic components. For patient E10, both the population interictal events and the local interictal events at contact ICL1 exhibited a frequency component around 2Hz (Fig. 3b). We conducted similar analyses on patients from the Yuquan and Epilepsiae datasets and performed significance testing on the extracted periodic frequencies after removing non-periodic components (see Methods). Most patients have representative electrodes exhibiting clear periodic characteristics (15/18 for the Yuquan dataset and 15/20 for the Epilepsiae dataset; Fig. 3c), and about half of the patients had population events that were also periodic (9/18 for the Yuquan dataset and 11/20 for the Epilepsiae dataset). As the occurrence of interictal events generally falls below 4Hz and the power spectral density (PSD) below 0.5Hz is difficult to fit accurately, the extraction of periodic frequencies was limited to 0.5-4Hz. The resulting periodic frequencies of interictal events in patients were mostly around 2Hz, as indicated by the interquartile range values on representative contacts (1.92 [1.75, 2.24]Hz for the Yuquan dataset and 1.94 [1.73, 2.36]Hz for the Epilepsiae dataset; Fig. 3c) or for population activities (1.76 [1.58, 2.21]Hz for the Yuquan dataset and 2.11 [1.82, 2.66]Hz for the Epilepsiae dataset; Fig. 3c).

In contrast to the periodicity of interictal events, where an event occurs every few hundred milliseconds, interictal events can also exhibit a certain randomness, remaining silent for several seconds before reoccurring (Supplementary Fig. 7a). This randomness might follow non-trivial regularities. We analyzed the time intervals between adjacent interictal events and found that both the population events of patient E10 and the local interictal events at contact ICL1 exhibited characteristics of a power-law distribution (Supplementary Fig. 7b). Power-law fitting of the interictal event interval distributions in the Yuquan and Epilepsiae datasets revealed that most patients’ population or local interictal event interval distributions exhibit power-law characteristics, as indicated by high fitting R^2^ values (Supplementary Fig. 7c). This power-law distribution of event intervals is also commonly reported in physiological neuronal discharges^49,50^ and might reflect similar underlying structures, noise, or dynamical properties.

**Fig. 7.**
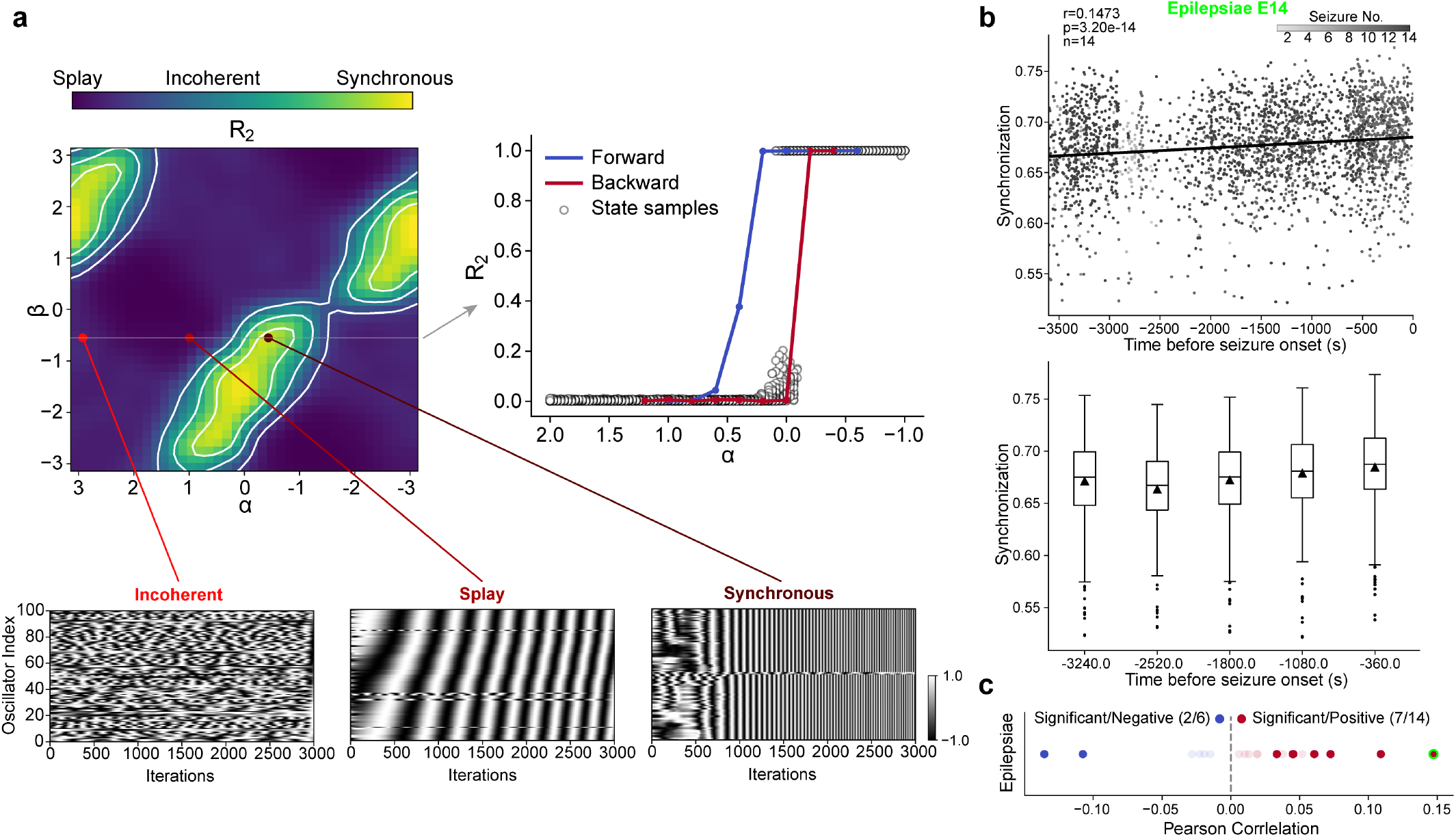
The transition from interictal to ictal states reflects a phase transition of network-level synchronization. **a**, The adaptive Kuramoto model can represent the ictal transition of epileptic networks. Depending on two structural parameters, it can exhibit three steady states - incoherent, splay, and synchronous -each characterized by a different level of synchronization (*R*_2_). For a certain type of plasticity (β fixed), the system state as a function of the interaction phase delay (α) exhibits hysteresis (critical bistability), indicating a first-order phase transition system. This suggests a common network mechanism underlying the interictal and ictal phases, with the transition from interictal to ictal reflecting a system phase transition toward greater synchronization. **b**, Prior to seizure onset, the synchronization level of interictal population events increased in patient E14. The middle panel provides a statistical summary of the above panel by segment, with the mean represented by a triangle. The bottom panel shows the preictal trend of synchronization of interictal propagation across Epilepsiae patients, showing a general upward trend. Patient E14 is marked with a green circle.

### 3. Hopfield Kuramoto Modelling of Interictal Propagation

The periodic occurrence of interictal population activities suggests that the SOZ functions as a dynamically coupled multi-node entity. This periodicity suggests that oscillator models, such as the Kuramoto model, can be used to characterize and study the dynamics of interictal network activities. The Kuramoto model is widely used to study synchronization phenomena in complex systems^51^. This approach aligns with the observed periodic patterns in epilepsy and offers a framework for understanding how different nodes within the epileptic network might synchronize their activities, contributing to the overall dynamics observed in epileptic seizures.

We have observed recurrent, stereotyped propagation during interictal periods, closely resembling the replay of neuronal discharges in memory consolidation^52–57^. Given the extensive presence of Hebbian-like plasticity in the brain^58,59^, it is reasonable to believe that epileptic networks might gradually form and solidify through the inter-node communication reflected in successive interictal propagation events. In the case of memory consolidation, Hopfield networks have been widely studied as models for encoding specific memories^60–63^ (Fig. 4a). Hopfield network encodes specific patterns into network connections using Hebbian rule. The stable propagation of interictal events is akin to the replay of specific memories in the epileptic network. We then employed the Hopfield model in conjunction with the Kuramoto model to study the encoding of specific propagation phase sequences in interictal data (Fig. 4a). In this approach, within the Kuramoto model, we used a Hebbian-like method to encode specific phase pattern that corresponds to the average interictal propagation sequence into the network structure of inter-node connections (Supplementary Fig. 8a), termed the interictal Hebb network (based on the cosine of the phase difference between nodes, see Methods for details). We found that the interictal Hebb network exhibits a linear recurrent structure related to the propagation sequence (Supplementary Fig. 8b,c). Supported by the interictal Hebb network, we were able to reproduce patient-specific, stable interictal propagation sequences (Fig. 4b). The propagation patterns obtained from the simulations were fully consistent with the actual interictal propagation sequences observed in the real data (Fig. 4c,d,e).

The symmetric nature of Hebbian rule ensures that the interictal Hebb network itself possesses characteristics of a real symmetric network structure. This implies that a Hebb network corresponding to a certain phase sequence also supports convergence to the exact opposite phase sequence (Fig. 5a,b). This type of symmetric plasticity mechanism has also been discovered physiologically^64,65^ and can explain the two exactly opposite interictal propagation patterns observed in patient E3. The initially unclear interictal propagation pattern of patient E3 became more distinct into two types of propagation patterns after clustering (Fig. 5c). Both categories of propagation patterns have corresponding real propagation activity samples (Fig. 5d), and their average propagation patterns show a significant negative correlation (Fig. 5e). These results suggest that the Hopfield-Kuramoto model can effectively explain stable, repeated interictal propagation sequences, including previously inexplicable coexisting opposite propagation patterns, supporting the involvement of plasticity mechanisms in the formation of epileptic networks. Interictal replay of propagation sequences consolidates the structure of the epileptic network, which in turn supports the retrieval of specific chronological ‘memories’. From the perspective of long-term plasticity, this reflects the important role of regular sequential activities in the formation and maintenance of epileptic network dynamics.

This model can also be utilized to explain the stochastic characteristics of interictal events. By imposing node-specific noise (modified Ornstein-Uhlenbeck Process noise, aka integration of Gaussian noise, see Methods for details) on the model, we were able to replicate the power-law distribution characteristics of the interictal event intervals observed in the data (Supplementary Fig. 7b; Supplementary Fig. 9b). In this process, due to the synchronizing effect of the Kuramoto model, the heterogeneous noises of the nodes are somewhat offset by the interactions between them, allowing the entire system to maintain synchronization and ultimately share a common velocity trend (the common velocity equals the mean of the node noises, see Methods) (Supplementary Fig. 9a). Driven by this common phase velocity, the distribution of the intervals (periods) of propagation activities exhibits power-law distribution characteristics similar to those in the data (Supplementary Fig. 9b). This suggests that specific types of noise activities might be involved in the occurrence of interictal events and that the SOZ as a whole might integrate and resist these local disturbances.

### 4. The Interictal Hebb Network Resembles Real Ictal Network

We conducted a joint analysis of the interictal Hebb network and seizure data to ascertain the physiological significance of the interictal Hebb network and its relationship with seizures. To construct the ictal network, we calculated the average coherence between pairs of iEEG contacts in the 1-250Hz range at the onset of seizures. We utilized Spearman correlation to evaluate its consistency with the interictal Hebb network (Fig. 6a-c). Our findings reveal a notable consistency between the interictal Hebb network and the ictal networks, indicating that the interictal Hebb network is a physiologically meaningful network. Additionally, it suggests that the interictal and ictal event phases share network structures to a certain extent (proportions of seizures that were significantly consistent with interictal networks: 0.65 [0.00-1.00] for Yuquan and 0.88 [0.29-1.00] for Epilepsiae, Median [25th quantile-75th quantile], FDR-corrected for each patient; Fig. 6d).

However, the results also show a certain degree of difference between interictal and ictal phase networks. Therefore, we further explored the changes in the consistency between interictal and ictal networks during seizures. We divided seizures into consecutive time windows and calculated network consistency within each window. By aligning multiple seizures based on their onset and termination times, we found that network consistency specifically increased at the onset of seizures (Fig. 6e) and dynamically varied throughout the seizure process. This suggests, on one hand, that the interictal Hebb network may contribute to the initiation of seizures by providing network support for early inter-node communication; on the other hand, it also indicates that there might be dynamic changes in network structures around seizure events, in contrast to the stable network structure supporting repeated propagation activities during the interictal periods.

### 5. Phase Transition from Interictal to Ictal States

The Hopfield-Kuramoto model unraveled the role of stable interictal network structures, formed under long-term plasticity, in the development and occurrence of epileptic activities^66– 68^. However, how do transient and unpredictable seizures occur? Previous work has reported the presence of short-term dynamic plasticity prior to seizures^69,70^. We thus hypothesize that short-term plasticity might lead to temporary changes in network structure, further driving the transition from interictal to ictal states. To explore the switching mechanisms between these two different epileptic network states, we introduced the capability for network structure to adaptively change with node states in the Kuramoto model (dynamic short-term plasticity), as opposed to the fixed network connections in the Kuramoto model or the Hopfield-Kuramoto model (network structure encoded before simulation, shaped by long-term plasticity, and relatively stable). This leads to a variant of the Kuramoto model, the adaptive Kuramoto model^47^, which incorporates real-time plasticity in the connections between oscillators.

Together with the Hopfield Kuramoto model mentioned above, we refer to them collectively as Kuramoto oscillator network with adaptive connections (KONWAC) model. This model also includes two main parameters: the phase delay *α* between node interactions and the parameter *β* that determines the type of plasticity, with *β* modulation allowing for different plasticity mechanisms like Hebbian rules or Spike-Timing-Dependent Plasticity (STDP) (see Methods for details). By varying these parameters, the system can produce states with varying degrees of synchronization (*R*_2_), including incoherent, splay, and synchronous states. The splay state approximates the stable propagation observed in the interictal phase, while the synchronous state, being more synchronized, approximates a more intense seizure state (Fig. 7a). Further examining the transition mechanisms between these states, we found that changing *α* while maintaining *β* could shift the system between splay and synchronous states. Monotonic changes in *α* in opposite directions lead to separated stable *R*_2_ curves, exhibiting a hysteresis phenomenon (Fig. 7a). This hysteresis suggests the system is a first-order phase transition system, showing bistability at a critical point. This aligns with previous beliefs that epilepsy is a first-order phase transition system^71,72^ and could partially explain the unpredictability of seizures, as noise might randomly push the system into different states. The lack of critical bistability in the original Kuramoto model also highlights the necessity of introducing network plasticity for studying epileptic state transition (Supplementary Fig. 10). Our analysis suggests that interictal propagation and seizures exist within the same dynamic network, and changes in structural parameters like communication efficiency (interaction delay α) can cause rapid changes in network topology, triggering a systemic phase transition from interictal to ictal phases at the network level.

To validate the hypothesis that seizures, as depicted in the above KONWAC model, result from a transition from a splay state to a more synchronized state, we examined whether there was an increasing trend in the synchronization level of interictal population activities before seizures in the Epilepsiae dataset (see Methods). We chose to use preictal activities rather than ictal activities, as ictal activities themselves are morphologically diverse and distinctly different from interictal activities. Our results indicate that the synchronization level of interictal activities tends to rise before the onset of seizures (14/20 patients showed increased synchronization, with 7/14 being significant, while 6/20 showed decreased synchronization, with 2/6 being significant; Fig. 7b,c). This finding supports the model’s prediction of increased synchronization during the ictal phase, suggesting that the state transition from interictal to ictal reflects a switch between types of different synchronization in the system. In summary, our study proposed and validated an adaptive complex model of the epilepsy network-KONWAC, dynamically unifying the interictal and ictal states.

## Discussion

Identifying the core network structure of epileptic networks and revealing the dynamic rules governing their spatiotemporal evolution will deepen our understanding of epilepsy mechanisms and pave the way for innovative diagnostic and therapeutic approaches. In this study, we analyzed two independent intracranial EEG datasets, totaling 4131.9 hours across 38 patients. We found that interictal discharges in the seizure onset zone (SOZ) exhibit a stereotyped spatiotemporal pattern, suggesting the presence of a stable core network structure within the SOZ. This core network structure is reinforced by regular interictal discharge activity through long-term plasticity, which, in turn, leads to the stereotyped spatiotemporal propagation sequence. The structure of the epileptic network and its neural activities co-evolve and mutually constrain, exemplifying the Hebbian principle in epileptic networks^73^. Contrast to the deterministic component in this process, under the influence of faster-scale dynamic plasticity, phase transitions of synchrony in the epileptic network drive the transition from interictal to seizure activity, which is more transient and fragile. Our proposed Kuramoto oscillator network model with dynamic plasticity in connections (KONWAC) unifies the deterministic core dynamics of epileptic networks and the stochastic seizure phenomena from a network dynamics perspective.

### The significance of interictal stereotyped propagation sequences in the diagnosis and treatment of epilepsy

In epilepsy surgery, the most crucial challenge lies in the precise localization of the SOZ. When using high-frequency activity for localization, distinguishing between physiological HFOs and pathological HFOs can be challenging, potentially leading to reduced specificity^74,75^. In this study, we observed synchronized activation of interictal events, suggesting that physiological high-frequency activity may not be a participant in this pathological population activity. This allows us to apply synchronization constraints to exclude interference from physiological HFOs or other high-frequency noise (Supplementary Fig. 3). Frequent co-activation also suggests exploring interictal events and their underlying network mechanisms from a holistic perspective^76–79^. With a deeper understanding of interictal events and their spatiotemporal propagation patterns, coupled with advances in data processing techniques, interictal data may better complement or even replace seizure data in clinical diagnosis and treatment.

Although the SOZ plays a crucial role in driving seizure propagation or interictal network activity^22,41,42^, it is often simplistically regarded in clinical practice as a homogeneous or somewhat messy region, deemed in need of complete resection. However, it may contain a wealth of overlooked details that could aid our understanding of epilepsy mechanisms. Our study unravels the presence of stereotyped interictal population events within the SOZ, suggesting a fine-grained network structure that supports temporal distinctions between different locations. These temporal sequences of interictal discharges serve as a valuable tool for elucidating the structure of the SOZ. For instance, it is particularly helpful in cases like insular epilepsy, which shares remarkably similar symptoms with temporal lobe epilepsy, making it challenging to infer the lesion’s location. Additionally, insular epilepsy itself may encompass a variety of seizure forms, the specific propagation pathways of which are yet to be determined^80^. By employing stable propagation sequences, it becomes possible to precisely localize the origin or specific propagation pathways of insular epilepsy, as illustrated in Supplementary Fig. 6, where two patients were confirmed to have similar propagation pathways originating in the insula and extending towards the frontal and cingulate gyrus. Insular epilepsy, a subtype of intra-temporal lobe epilepsy, highlights the applicability of this interictal chronology approach to different focal epilepsy types, including both intra- and extra-temporal lobe epilepsy. In the Yuquan dataset, primarily consisting of patients with FCD, TSC, and other focal epilepsies, stable propagation patterns generally originated from localized cortical structures outside the temporal lobe. In contrast, the Epilepsiae dataset, primarily comprising medial temporal lobe epilepsy patients, exhibited stable propagation patterns originating from the temporal lobe or the hippocampus, which, due to its position in functional circuits like the Papez loop, may have broader global brain network effects^20,81^. Across these different epilepsy types in the two datasets, stable interictal propagation sequences and their origins in the brain could be readily identified (Supplementary Fig. 4; Supplementary Fig. 6b). These findings demonstrate the potential of using interictal data to complement or even substitute seizure data in clinical diagnosis and treatment, thanks to advanced data processing techniques and a deeper understanding of interictal events and their spatiotemporal propagation patterns.

### Deterministic components in epileptic networks

In general, epilepsy is often perceived as a disorder filled with randomness due to the unpredictability of seizures. However, it is known that *scattered braves cannot form a formidable army*, and there must be regular components involved in the formation and maintenance of epileptic networks. In this study, we observed periodicity in the occurrence of interictal events, suggesting the existence of microcircuit structures near iEEG electrodes that support periodic activity^82–85^. In this study, the periodicity of interictal events suggests that we can abstractly represent local activity using oscillator models, and the interactions among multiple oscillators can be characterized using the Kuramoto model to describe the overall SOZ network. The Kuramoto model, as a dynamic system for multiple oscillator coupling, has been widely used to study synchronization phenomena in complex network systems^86^. Currently, there are efforts to apply it to the study of the mechanisms underlying epileptic seizures, suggesting that seizures may correspond to explosive synchronization of the system^87^. Moreover, the periodic interictal synchronous discharges, characterized by repetitive propagation, suggest that the SOZ is a dynamic entity composed of multiple coupled nodes under specific structural constraint, and the Kuramoto model is evidently a concise dynamic model capable of characterizing the essentially synchronous epileptic networks.

Another deterministic phenomenon of epilepsy is the stable interictal propagation pattern within the SOZ, which reflects the presence of a stable driving network structure. However, the repetitive propagation sequence itself does not inform us about the specific network structure or driving mechanism within the SOZ. Although simple chain-like structures such as synfire-chain can generate highly stereotyped repetitive propagation patterns, this model requires stimulation at the head of the chain^79,88^ and cannot account for certain random variations in the propagation patterns beyond their repetition. The SOZ network supporting interictal repetitive propagative activity appears to have a more specialized structure. We propose that stable interictal propagation closely resembles memory replay during sleep, much as repetitive replay enhances memory consolidation under conditions of brain plasticity^64,89^. Such interictal repetitive discharges may play a critical role in the formation and consolidation of epileptic networks.

In studies concerning the encoding and storage of memories, Hopfield networks have been widely employed. They use Hebbian plasticity rule to encode, store, and retrieve specific activity patterns^60,61,90^. Unlike traditional Hopfield networks that encode discrete signal patterns, stable interictal propagation activities are essentially patterns of temporal order (phase patterns). Combining Hopfield networks with the Kuramoto model allows for the encoding of specific phase patterns. This combination uses a Hebbian-like rule to store the network structure within the Kuramoto model^46,91^, where specific patterns of node activity determine inter-node connectivity. The network formed by this encoding mechanism, in turn, supports the reproduction of propagation patterns and can explain previously unexplained phenomena of a pair of opposite propagation patterns^22^ (Fig. 5). This is essentially a coupling or mutual constraint between activity and connections, where activity, under plasticity conditions, promotes connection patterns, and connection patterns simultaneously support and constrain the propagation of activity, i.e. an idea of coevolution^92^. This intertwining of neural activity and network structure is manifested in the formation of epileptic networks. For instance, some work has shown that as epilepsy progresses, there is adaptive strengthening of myelin sheath within the epileptic focus^66^. Corresponding to the formation of the epileptic network, this also inspires the idea that we can use electrical stimulation to perturb network activity, thereby altering the network structure and subsequently affecting the overall pathological manifestations of epilepsy. Related work with responsive neural stimulation (RNS) has already indicated that long-term electrical stimulation achieves therapeutic effects in epilepsy primarily through network reconstruction^67^. Subsequent studies monitoring the early stages of epileptic network formation or observing the effects of stimulation on network physiology will further validate our theory.

Certainly, we must also recognize that periodic local activity, in conjunction with stable plastic propagation behavior, collectively contributes to the formation of epileptic networks. However, compared to the latter’s general role in brain function, the former may hold more specific significance in the formation of epileptic networks. If local activity lacks periodicity and instead occurs randomly, it would be impossible for different nodes to establish fixed connections, leading to the inability to form a stable core structure within the epileptic network. Periodic local activity may be intrinsic to the formation of epileptic networks, prompting us to further investigate the microcircuitry structures or local excitatory/inhibitatory imbalance mechanisms^93,94^.

### Random components in epileptic networks

Epilepsy is most notorious for its unpredictability, which brings significant inconvenience and danger to the life and safety of patients^95^. It has also given rise to clinical issues such as epilepsy prediction and feedback control devices such as RNS^96^. Deciphering and understanding the sources of randomness in epilepsy will allow us to better control epilepsy and benefit patients.

Previous research has shown that there are short-term plasticity changes preceding seizures, such as NMDA-related EEG activity alterations^69^ or accelerated interictal activity propagation before a seizure^70^. This supports the use of an adaptive Kuramoto model with short-term plasticity as a concise epilepsy network model for studying the transition mechanism from interictal to ictal states. We found that changes in phase delays in the adaptive Kuramoto model’s node interactions lead to a phase transition in the collective behavior of the system, shifting it from a propagating state to a more synchronized state. These phase delay changes may arise from an activity-dependent shift in the overall neurotransmitter levels within the network^97^ or perhaps rhythmic fluctuations of physiological factors, such as expression of ion channels and neurotransmitter receptors^98,99^.

Previous work has, to some extent, confirmed the view that “the interictal and ictal periods correspond to two steady states of epilepsy, and their mutual switching corresponds to the phase transition of the system caused by changes in certain structural parameters”^71,72,100,101^. For example, decreased resistance to perturbations before a seizure ^71^ or the so-called critical slowing-down phenomenon ^72^ suggests that the network before a seizure is in a critical state between different states and is more sensitive to various noise perturbations. However, unlike the previous view that the phase transition originates from local nodes, our model and empirical findings suggest that seizures may stem from a phase transition in the collective behavior of the network. In this process, a seizure reflects a more synchronized state of the epileptic network (Fig. 7). Moreover, the coevolution of network structure and dynamics imparts a positive feedback effect on the changes in network activity, endowing the adaptive Kuramoto model with the characteristics of a first-order phase transition system, wherein the system state can undergo a sudden change at the critical point^102,103^. This aligns with the consensus from previous physiological and modeling work that seizures are consistent with being a first-order phase transition system^71,72^. The bistable nature of first-order phase transition systems at the critical point allows random noise to stochastically push the system across state boundaries, making seizures difficult to predict. Since in coevolving systems, both node dynamics and network structure collectively determine the overall state of the network, system state noise related to both these aspects may stem from fluctuations in molecular mechanisms such as internal ion concentration gradients or neurotransmitter levels^104,105^ or may be influenced by external environmental or behavioral factors^106,107^.

In summary, compared to the traditional Hopfield-Kuramoto model, which replicates deterministic interictal activities under long-term plasticity in the construction of epileptic networks^66,68^, the adaptive Kuramoto model explains the random transition to seizure caused by rapid changes in network topology under short-term plasticity. The KONWAC model we proposed essentially combines two scales of plasticity, shaping both the stereotyped propagation and the randomness of seizures in epileptic networks through the interplay of regularity and uncertainty.

### Personalized epilepsy modeling based on long-term interictal data

With the advancements in data and computational power, our proposed KONWAC epilepsy dynamics model can enable individualized parameter modeling and serve as a comprehensive prior for clinical applications. In this regard, initiatives like virtual epileptic patient (VEP) utilize multimodal patient data to construct personalized seizure propagation models. These models are applied to clinical needs such as lesion localization and surgical guidance through parameter inference and virtual resections^6,108–110^. Other work focuses on guiding the optimization of parameters for epilepsy stimulation control by constructing thalamo-cortical models and similar approaches^111,112^. These efforts provide examples of the application of computational models in clinical settings and point towards a new direction in epilepsy clinical research.

We should note that in the modeling of epilepsy network dynamics, abundant data can better help uncover deterministic components within pathological underpinnings, avoiding bias introduced by stochastic components. Seizure data, due to their short monitoring duration, may not always guarantee reliability in modeling tasks. In contrast, interictal data, which are easier to obtain than seizure data, more directly represent the dynamics of epilepsy network evolution and may reflect the more fundamental dynamic mechanisms of the epilepsy network as evidenced in current study.

As our understanding of the role of long-term plasticity mechanisms in epilepsy treatment and the technical development of closed-loop control of epilepsy continues to grow^113–115^, using interictal data for personalized modeling is better suited to such emerging trends. Personalized epilepsy network modeling holds promise for establishing new control strategies, especially those based on the aforementioned fast and slow plasticity mechanisms for electrical stimulation control.

## Methods

### Yuquan Dataset

#### Patients

This study utilized a dataset from 18 patients with refractory epilepsy treated at Tsinghua University’s Yuquan Hospital Epilepsy Center (see Supplementary Table 1 for patient details). All underwent SEEG electrode implantation for in-depth diagnosis. Data use was authorized by the hospital, focusing mainly on patients with focal etiologies like FCD and TSC. All participants, or their legal guardians, provided informed consent to participate in this study.

#### SEEG Data

Continuous SEEG data were collected in the hospital using NIHON KOHDEN EEG-1200C amplifiers at 2 kHz. Electrodes, 2 mm long with a 3.5 mm gap, targeted lesions in cortical and subcortical areas as clinically required. Patients provided at least 24 hours of SEEG, including multiple seizures. If initial data lacked seizures, additional episodes were added for comprehensive study of interictal and ictal phases.

#### SEEG Preprocessing

The preprocessing of SEEG data involved several steps: removing contacts with significant noise, resampling to 800Hz, and applying a 50Hz notch filter (second-order IIR, quality factor of 30^116^). A bipolar re-referencing method was used, taking the difference between adjacent contacts and retaining the inner contact’s name. Seizure data were processed similarly and trimmed to equal lengths.

#### Image Data and Processing

For each patient, the image data include preoperative T1 MRI and postoperative CT scans. Cortical reconstruction was carried out on MRI images via Freesurfer^117^. MRI and CT images were aligned using FSL^118^, facilitating manual marking of electrode coordinates.

### Epilepsiae Dataset

#### Patients

This study includes 20 refractory epilepsy patients from the Epilepsiae dataset^45^ (see Supplementary Table 2 for patient details). Most provided at least 100 hours of continuous intracranial EEG, with extensive clinical annotations such as seizures, diagnoses, and surgical outcomes. Out of 30 patients in the Epilepsiae dataset, 20 were chosen based on the sampling rate criteria. Data use was authorized by the custodian.

#### Intracranial EEG Data

The Epilepsiae dataset includes patients with varying EEG sampling rates, with a minimum of 512Hz, and includes diverse electrode configurations such as electrode strips, electrode grids, and stereo-electrodes, positioned according to clinical needs. *Intracranial EEG preprocessing*. The preprocessing of EEG data is consistent with that of the Yuquan dataset, except that a global average re-referencing is used, due to the diversity of electrode configuration.

#### Image Data and Processing

For each patient, the image data includes a skull-stripped T1 MRI and electrode site voxel positions within the MRI. Cortical surfaces were reconstructed using Freesurfer. Electrode site voxel positions were then mapped to cortical space coordinates using the MRI’s coordinate transformation matrix.

### Interictal events detection and collective population activity

To detect IEs, we filtered EEG data with an 80-250Hz band-pass filter (fifth-order Butterworth) and extracted the energy envelope via Hilbert transform. We noted energy surges within this band during ripples or broadband spikes. A threshold was established to pinpoint these high energy instances, aiding in IE identification:

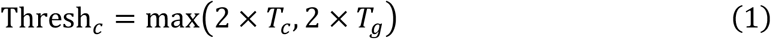

Here, *c* represents the contact index, with *T*_*c*_ and *T*_*g*_ representing the median envelope values for contact *c* and all contacts, respectively. We optimized our event detection with post-processing, merging windows less than 20ms apart to prevent duplicates, selecting windows of 50-200ms, and ensuring the window’s average energy is at least 1.5 times that of its surroundings, consistent with IE characteristics. To expedite the main detection steps (filtering, envelope extraction, detection, etc.), we employed Python packages such as cupy and cusignal^119,120^, which utilize GPU acceleration for signal processing (as shown in Supplementary Fig. 11).

We noted that interictal events frequently occur in sync, underscoring a compact cluster of abnormalities in time and space. This synchrony helps filter out noise or physiological high frequencies, leading us to introduce a two-step method as shown in Supplementary Fig. 12.

In the first step, we enumerated IEs at each site to locate the SOZ. We then mapped all IE timings onto a unified time axis, creating an IE occurrence rate curve. Population events were identified by thresholding periods of elevated IE activity. Reinforced by the above synchrony constraint, alienated noisy events can be filtered out. In the second step, reevaluating IEs within these population events further clarified the SOZ’s location. We relocated the final SOZ nodes and their associated population events, which informed our subsequent interictal network analysis. We employed the Area Under the Curve (AUC) metric to assess the consistency of IE counts with the clinical SOZ across the two steps.

To address the high synchronization of IEs within the SOZ, we calculated the number of overlapping IEs between pairs of sites, both within and outside the SOZ. This process resulted in two matrices: one representing synchronized IEs within the SOZ, and another for those outside the SOZ. The mean of each matrix was used to evaluate the difference in IE synchronization between the inside and outside areas. For consistent comparison across patients, the matrices for each patient were normalized by the highest common value found in these two matrices.

### Propagation order characterization and test of pattern repeatability

To characterize the network propagation features of IEs during population events, we utilized the centroid of the time-frequency spectrum as a reference point. This was based on the presence of island-like high-frequency components in the HFO time-frequency graph. The signal was filtered into the ripple band of 80-250Hz, and then the spectrogram *S* ∈ ℝ^*T*×*F*^, was calculated, where *T* represents the number of time samples and *F* represents the frequency samples. The matrix *S* was normalized to ensure that its sum equals 1, serving as a form of weighting. By utilizing the spectrogram centroid formulas provided below,

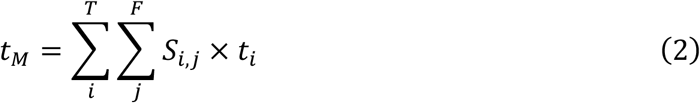

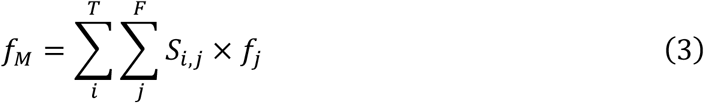

we calculated the time and frequency of the centroid, where *i* is the time index, *j* is the frequency index, and *t*_*i*_ and *f*_*j*_ represent the corresponding time and frequency, respectively. Utilizing the centroid time, we characterized inter-site transmission during population events. Sorting the time sequence enabled us to ascertain the rank of each site and the propagation pattern.

To assess the repeatability of propagation patterns, where greater repeatability is indicated by smaller deviations from the average pattern, we computed an average pattern and compared each propagation pattern to it.

We compute the weighted average rank for each site, based on its three most frequent ranks in all propagation events. Sorting these average ranks yielded the average propagation pattern. To assess consistency between individual samples and the average pattern, we utilized the Matching Index (MI) method^22^, which measures the fraction of consistent relationship pairs between two sequences,

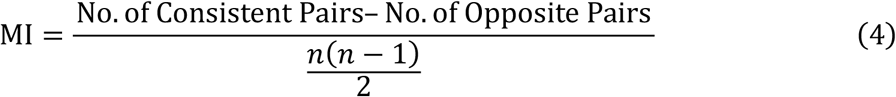

where *n* is the length of one sequence. An MI of 1 signifies complete consistency between the two sequences, whereas an MI of -1 indicates completely opposite propagations. We compared each individual propagation sequence to the average propagation sequence to generate a distribution of MIs. The more pronounced the repetitive propagation pattern is, the more right-skewed the overall MI distribution will be, and consequently, the median value of the MI distribution will be higher.

To test for the presence of repeatability in the propagation pattern, we conducted a permutation test. This involved randomly perturbing the propagation pattern of each population activity and then recalculating the median value of the Matching Index (MI) distribution. This perturbation process was repeated 200 times to generate a null distribution of the MI median values. The presence of repeatability in the propagation order was determined based on whether the actual MI median fell within the upper 5% (right-tail) of the null distribution, which corresponds to a significance level of p<0.05 (as illustrated in Fig. 2b).

Several spatiotemporal and spectral properties of interictal propagation were examined. Firstly, the propagation delay between two temporally adjacent sites was quantified. Secondly, the centroid frequency distribution was determined. Finally, to assess the spatial properties, we summarized the distribution of the upstream electrode contacts (the first 50% in the average propagation pattern) across different brain regions. These regions included the frontal lobe, insula, cingulate gyrus, and limbic system (encompassing the amygdala and hippocampus), among others.

### Fitting of periodicity and randomness components in interictal event occurrences

To investigate the temporal characteristics of interictal events, we initially transformed the detection results of these events into discrete event signals. Time windows that exceeded the threshold during the detection process were assigned a high signal value of 1, while all other times were assigned a low signal value of 0. A similar approach was adopted for population events as depicted in Fig. 3. Utilizing these derived discrete event signals, we conducted an analysis to identify the periodic and random components.

If the occurrence of interictal events is periodic, the Power Spectral Density (PSD) of these discrete event signals will exhibit a noticeable peak. To analyze this, we utilized the ‘fooof’ method^121^, which is designed to fit and decompose the PSD into its periodic and non-periodic components. The decomposition formula used in this process is as follows:

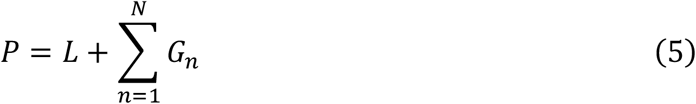

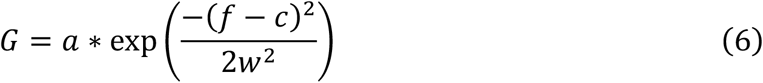

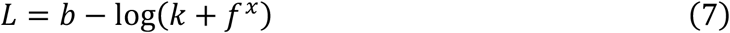

where *P* is the logged power values, and *f* is the frequency. *G*_*n*_ is a specific periodic component, where *a, c*, and *w* correspond to the energy, frequency, and bandwidth of this periodic component, respectively. *L* is the aperiodic component, where *b* and *x* are the compensation and power law exponent, respectively, and *k* controls the relative inflection point position within the aperiodic component.

To test for the presence of periodicity in the occurrence of interictal events, we utilized a t-test to determine whether the energy of the periodic frequencies found in spectral decomposition is significant, after removing the non-periodic components (Supplementary Fig. 13).

For the analysis of the random component within IE occurrence, we focused on the distribution of time intervals between adjacent interictal events, as indicated by the red double-arrowed lines in Supplementary Fig. 7a. The the distribution of inter-event intervals was observed to exhibit a power-law characteristic, a phenomenon also noted in the distribution of neuron firing intervals^49^. To perform a power-law fit for the distribution of time intervals (log-log scale), the following formula is used:

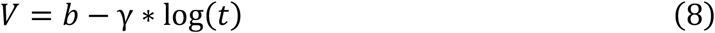

Where *V* is the logarithmic count, *γ* is the fitted power-law exponent, and *t* is the time interval between adjacent events. The fitted r-square value can be used to assess the quality of the fit.

### Hopfield Kuramoto Model

Based on the periodicity of interictal events, we can consider adopting the Kuramoto oscillator model to model the SOZ network^86^. The Kuramoto model is commonly used to study synchronization phenomena in weakly coupled complex networks:

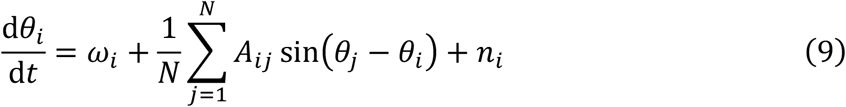

In this model, *θ*_*i*_ denotes the phase of node *i*, while *ω*_*i*_ denotes the natural angular frequency of node *i. A*_*ij*_ stands for the connection strength between nodes *i* and *j*, and *n*_*i*_ is the noise of node *i*. The noise *n*_*i*_ is modeled using the Ornstein-Uhlenbeck process (OUP)^49^, which is characterized by relatively slow variations and is commonly employed in the study of the randomness of neuronal discharges. The representation of OUP noise is as follows:

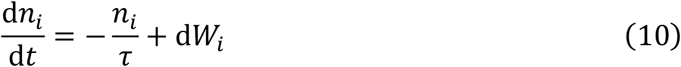

Here, *τ* represents the time constant of the mean-reversion term, where *dW*_*i*_ represents sampling from either a Gaussian distribution or a Pareto distribution.

We hypothesized that the development of epileptic networks is attributed to recurrent interictal propagation activity, suggesting a plasticity mechanism akin to Hebbian learning. This prompted us to transition from the Kuramoto model to the Hopfield-Kuramoto model^46^, which enables the encoding of phase patterns into the network structure. In the traditional Hopfield model, where the Hebbian rule emphasizes the establishment of connections between synchronously firing neurons, encoding is accomplished through the outer product of pattern vectors. For the Kuramoto model, the intuitive form of Hebbian rule states that the smaller the differences between nodes in the pattern vector, the stronger the connection. This can be realized by *A*_*ij*_ = cos(*ϕ*_*i*_ − *ϕ*_*j*_), where ***Φ*** = [*ϕ*_*1*_, *ϕ*_*2*_, …, *ϕ*_*N*_] is the interictal phase vector and *N* is the number of network nodes. On the other hand, the fact that a connection matrix encodes a particular pattern vector is equivalent to the system being able to converge to the propagation pattern under the influence of that matrix. To achieve this, some work in complex number field suggests that *e*^i*Φ*^ should act as an eigenvector of a connection matrix ***A***^*c*^, granting ***A***^*c*^ a form analogous to the outer product structure of the Hopfield model^122^, i.e.,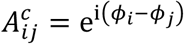. Taking the real part also yields *A*_*ij*_ = cos(*ϕ*_*i*_ − *ϕ*_*j*_) as the interictal Hebbian network structure.

In the simulation process, the transformation formula from the propagation pattern of the patient data [rank_1_, rank_2_, …, rank_N_] to the phase vector in the model [*ϕ*_*1*_, *ϕ*_*2*_, …, *ϕ*_*N*_] is as follows:

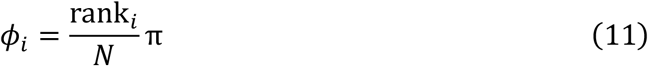

To validate that both forward and backward propagation patterns can emerge within the same network. Initially, we configured a pattern vector and derived the corresponding Hebb network structure. By maintaining this Hebb network structure constant and randomizing the system’s initial values on several occasions, we observed that the unaltered network structure could lead the system to converge to two diametrically opposite propagation patterns. In the analysis of patient E3’s data, we clustered the interictal propagation patterns into two categories and identified a pair of inverse propagation patterns, aligning perfectly with the predictions of our model (as shown in Fig. 5).

### Ictal coherence network

To assess the consistency of the network between the interictal and ictal phases, we computed the coherence network structure during the ictal phase. Coherence characterizes the stability of the phase relationship between two time series, namely the closeness of the two in the phase space. Considering a segment of signals synchronously recorded from two channels, *x* and *y*, we posited that this signal segment is divided into *n* parts. Consequently, the coherence at frequency *f* between these two channels within this signal segment is defined as follows^123^:

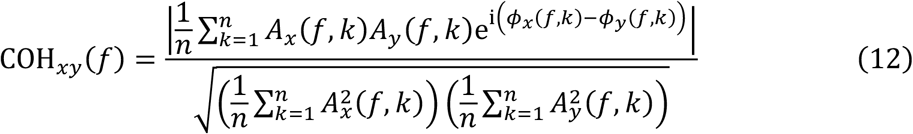

In this context, *A*_*x*_(*f, k*) represents the amplitude of the *f-*frequency component in the PSD of the *k*th part of the *x* signal. Similarly, *ϕ*_*x*_(*f, k*) denotes the corresponding phase. The coherence connection between two nodes in the ictal network was characterized by calculating the average coherence across the frequency range of 1-250Hz.

### Adaptive Kuramoto modelling of epileptic network

Given the presence of short-term plasticity prior to seizure onset^69,70^, we contemplated incorporating dynamic plasticity into the Kuramoto model to examine the transition mechanism between interictal and ictal states. This dynamic plasticity enables real-time coupling between the network connections and node dynamics, leading to a variant known as the adaptive Kuramoto model^47^. In this model, network connections and node dynamics are interdependently coupled, creating a dynamic entity. The Hopfield-Kuramoto model mentioned earlier, along with the adaptive Kuramoto model characterized by short-term plasticity, collectively constitute the KONWAC model. This integration is a significant enhancement, addressing the often-neglected aspect of physiological network plasticity in dynamic epilepsy modeling. The dynamic interplay between oscillator activity and connection strength is mathematically expressed as follows:

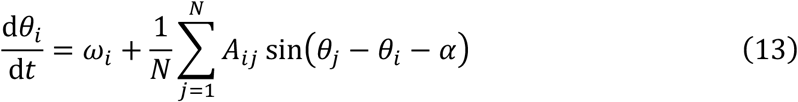

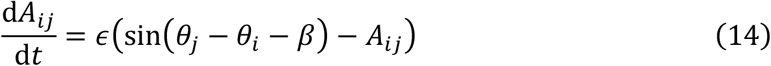

where *θ*_*i*_ and *θ*_*j*_ are the phases of two oscillators, *ω*_*i*_ is the angular frequency of oscillator *i, A*_*ij*_ is the coupling strength between oscillator *j* and *i*, and *N* is the total number of oscillators. *α* represents the delay between oscillators and *β* is a parameter that can set different modes of network plasticity, making the plasticity STDP-like or Hebbian-like. We set the angular frequency of the oscillator to follow a normal distribution, with the initial phase uniformly sampled from *0* − 2π, and the initial value of *A*_*ij*_ uniformly sampled from the range of *0* − 0.001.

In practical terms, plasticity can encompass both long-term and short-term components. The long-term component, which corresponds to the network connections in the Hopfield-Kuramoto model, changes gradually and is primarily influenced by regular factors. This component provides stability and maintains repetitive sequences of interictal propagation. Conversely, short-term plasticity exhibits more rapid fluctuations and is predominantly influenced by stochastic elements, resulting in more pronounced changes in network activity. Our study specifically focuses on the impact of short-term plasticity during seizure transitions.

### Bifurcation analysis of adaptive Kuramoto model

For the bifurcation analysis, we performed a parameter sweep of the phase delay *α* and the plasticity parameter *β* in the range from 0 to 2*π*. We used 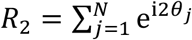 to evaluate the degree of synchronization in the final steady state under different parameter combinations and generated the topographic map depicted in Fig. 7a, where *θ*_*j*_ represents the phase of the *j*th oscillator and *N* is the number of oscillators.

To investigate the transition mechanism between interictal and ictal phases, we studied the phase transition law of the system between the splay state (similar to propagating IEs) and the synchronous state (similar to seizures). Pathological phenomena such as the preictal increase in IE propagation velocity^70^ may suggest preictal changes in network structural parameters related to propagation efficiency, which correspond to changes in the phase delay *α* between nodes in our model. Therefore, we fixed the parameter *β* and randomized the system multiple times under different *α* values to sample the bifurcation space of the system. We also monotonically increased or decreased *α* and recorded the corresponding steady state *R*_2_ values as *α* changed. As shown in Fig. 7a, the phase transition of the system between the splay and synchronous states is a first-order bifurcation with a critical bistability characteristic.

### Synchronization trend of IE propagation activity before seizure

To validate the adaptive Kuramoto model’s explanation of the transition between interictal and ictal phases, our objective was to demonstrate that seizures exhibit heightened network synchronization, as predicted by the model. However, given the diversity and irregularity of ictal activity patterns, our investigation focused on determining whether there was a discernible trend towards increased synchronization during preictal IE propagation activity. Given the previous research evidencing an interictal-to-ictal transition up to one hour prior to seizure onset^124^, we analyzed the synchronization trends in IEs during this critical one-hour preictal period. To quantify this, we employed the synchronization index (SI) to describe the degree of synchronization in each population activity,

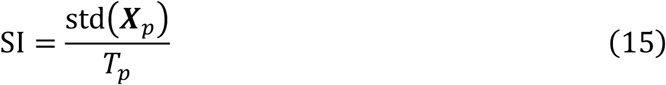

where ***X***_***p***_ ∈ ℝ^*C*^ represents the centroid moment vector of IE propagation activity, *C* is the number of sites, *stdi*(*⋅*) calculates the variance, and *T*_*p*_ represents the propagation time between the first and last sites. We kept only the seizures that occurred more than 3 hours after the preceding seizure, aligned the IE propagation activities within the one-hour period preceding seizures by the seizure onset time, and used Pearson correlation to examine the trend of synchronization in the propagation activity before seizures.

Using this method of measuring synchrony, we can see that the sequence [1,3,3,3,5] has stronger synchronization than the sequence [1,2,3,4,5]. In addition, the synchronization of the sequence [1,2,3,4,5] is equal to that of the sequence [0.1,0.2,0.3,0.4,0.5], thus eliminating the influence of propagation speed and focusing solely on the level of synchrony.

## Data Availability

All data produced in the present study are available upon reasonable request to the authors.

