## supplementary materials for "Replay of Interictal Sequential Activity Shapes the Epileptic Network Dynamics"

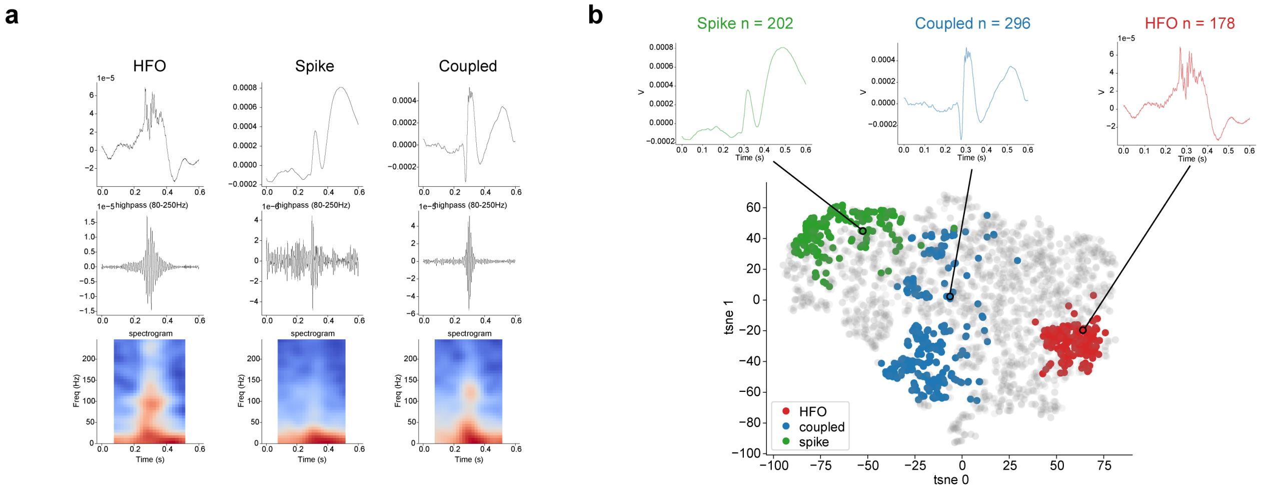


**Supplementary Fig. 1 | The three representative categories of interictal events. a**, The three representative types of interictal events are High-Frequency Oscillations (HFOs), spikes, and a coupled type where HFOs ride on spikes. **b**, The three categories of samples, manually selected from the interictal events of Yuquan patient Y1, can serve as representatives to construct a low-dimensional space distribution of the patient's interictal events. The colored dots represent these manually labelled samples, while the gray dots represent a total of 2600 samples collected from 26 contacts for the same patient. The low-dimensional features were obtained by applying the t-SNE algorithm to reduce the dimensionality of the original sample signals.

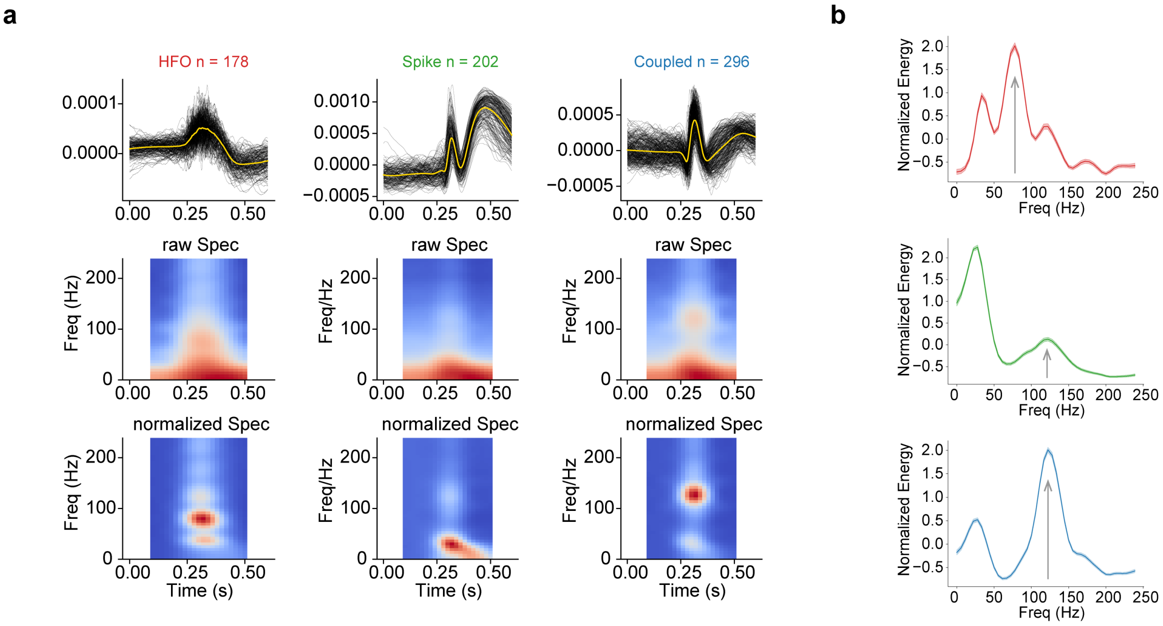


**Supplementary Fig. 2 | The time-frequency spectral characteristics of the three representative sample categories. a**, The average time-frequency spectrogram characteristics of the three sample categories, both in their original form and baseline-normalized form, with normalization performed using the mean of the first 0.15 seconds for calibration. **b**, The spectral energy distribution obtained from the temporal averaging of the middle 0.2 seconds in the normalized time-frequency spectrogram reveals specific energy enhancement in the high-frequency (>50Hz) range for all three representative sample categories.

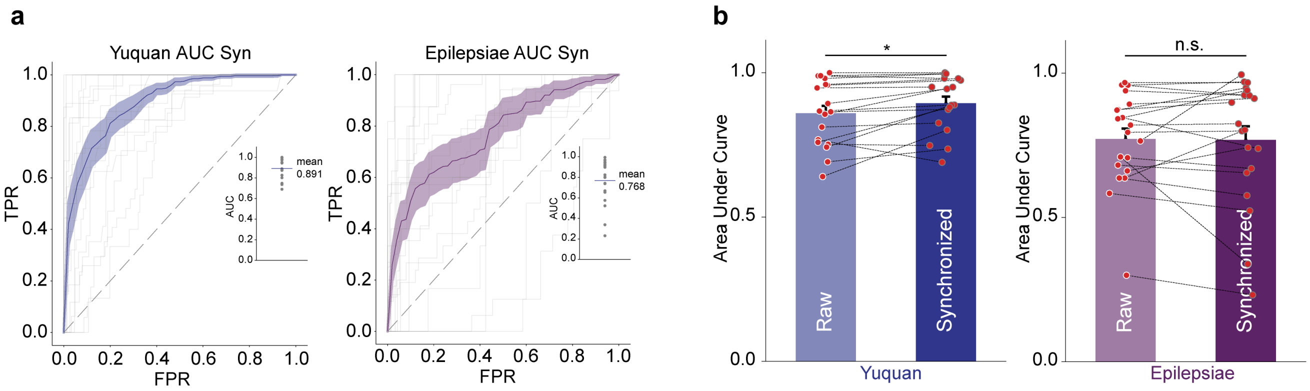


**Supplementary Fig. 3 | Influence of synchronicity of interictal events on SOZ prediction. a**, IE distribution with synchronicity constraint predicts the clinical SOZ well (AUC values obtained by ROC curve method are shown in the inset). The colored curve and shaded area represent the mean and s.e.m., respectively, while the horizontal line in the inset represents the mean AUC. **b**, Synchronicity constraint of IEs significantly improves the prediction of SOZ in the Yuquan dataset. Mean values are shown as bars, with s.e.m. shown as error bars.

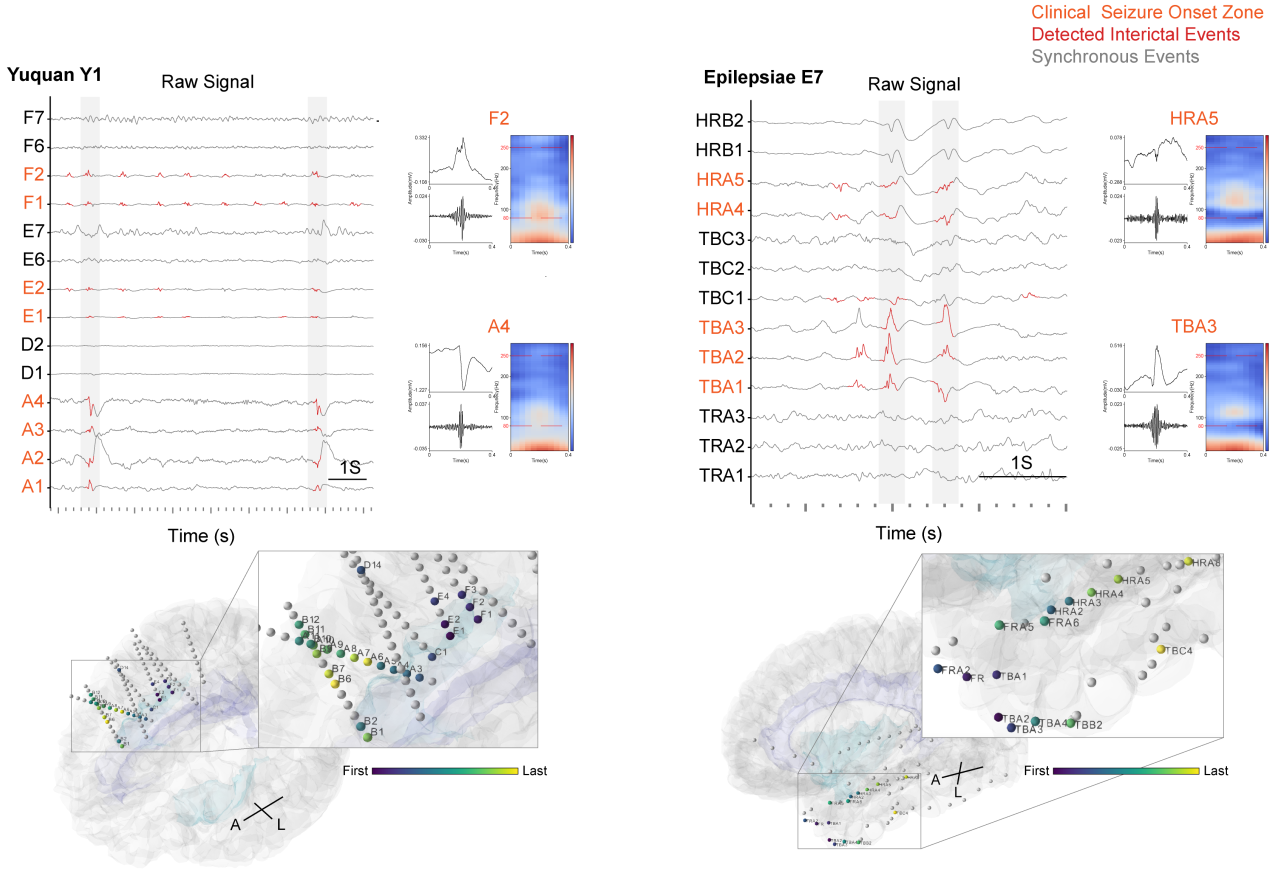


**Supplementary Fig. 4 | Interictal data of representative patients from two datasets.** Raw iEEG signal of patients in Fig. 1. Examples of detected interictal events are shown on the right with raw signal, 80-250Hz band-pass signal, and time-frequency map. The average propagation patterns of interictal events are projected onto electrodes and displayed below, showing two patients respectively exhibiting propagation from the insula to the frontal lobe and cingulate gyrus, and from the anterior to the posterior temporal regions.

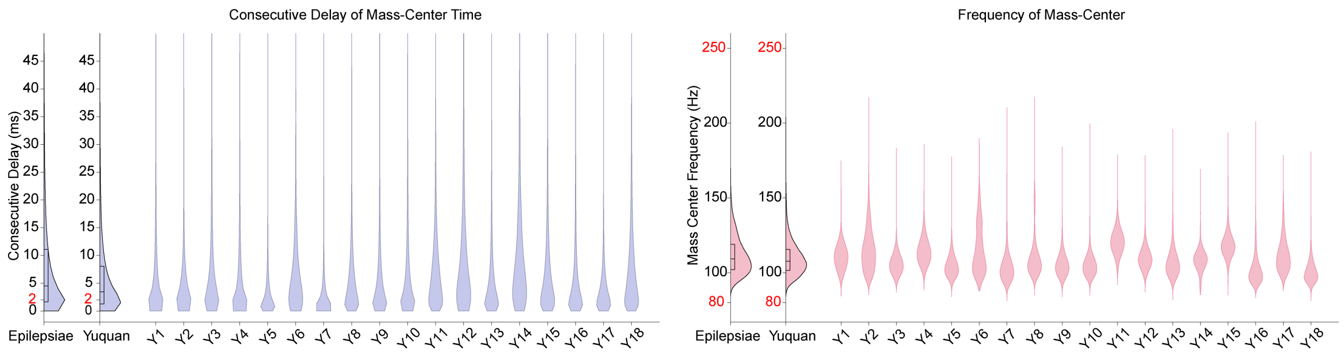


**Supplementary Fig. 5 | Temporal and spectral characteristics of interictal propagation.** The distribution of interictal event propagation delay between temporally consecutive contacts, as well as the distribution of centroid frequencies. The average propagation delay was about 2ms at the synaptic level. The overall distribution of the dataset was obtained from equal sampling of each patient, and only detailed results from the Yuquan dataset are presented.

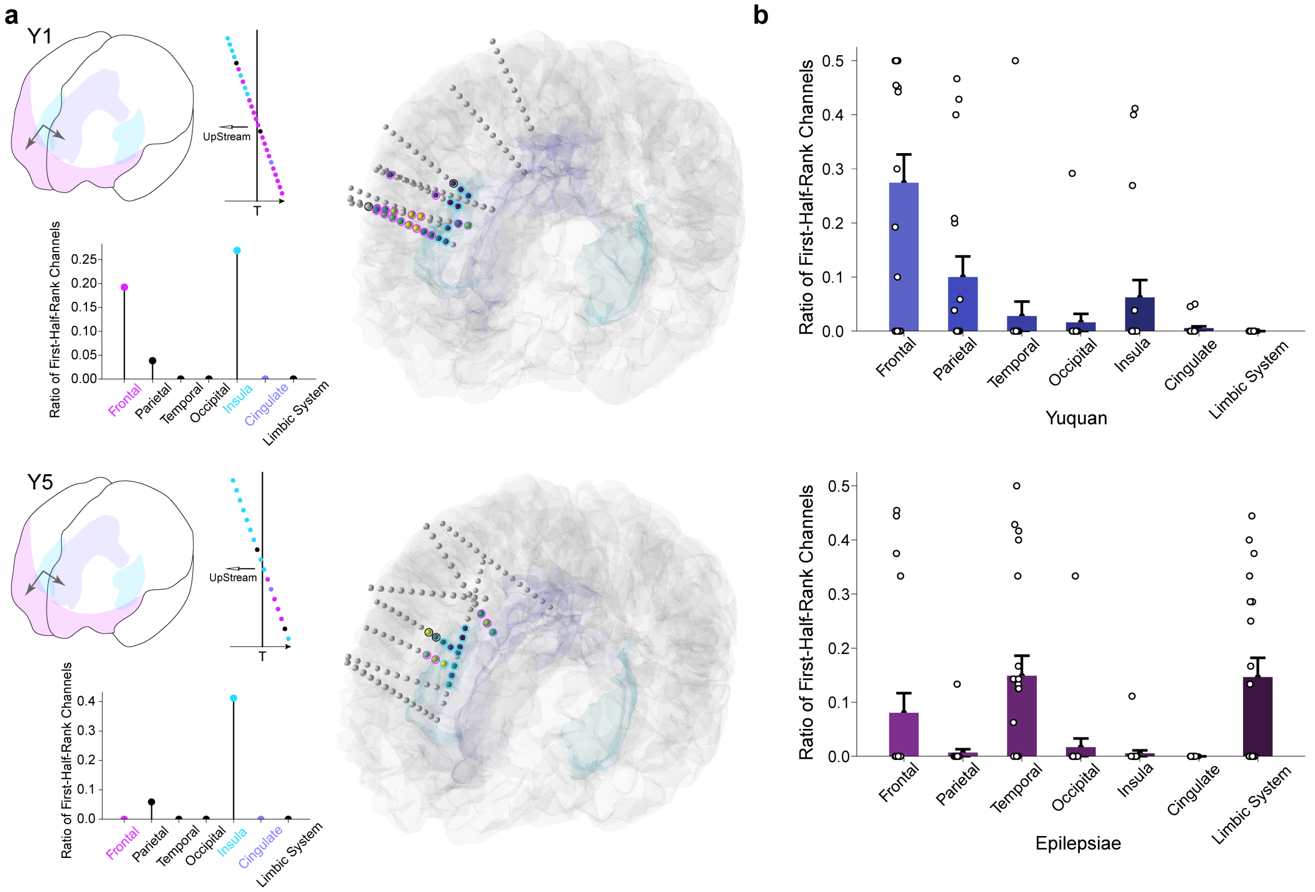


**Supplementary Fig. 6 | Stable interictal propagation shows the fine structure of SOZ. a**, In two patients with lesions involving the insula, cingulate gyrus, and frontal lobe, a common origin in the insula and spread to the frontal and cingulate gyrus was observed. The distribution of upstream electrodes in these three brain regions also indicated an insular origin. **b**, Distribution of origin sites of interictal propagation patterns in Yuquan and Epilepsiae patients. Similar to panel **a**, the distribution of each patient's upstream contacts in each brain region is summarized. In the Yuquan dataset, the majority of patients have FCD or TSC etiologies, and the origin brain regions involved are mainly located in the insula, frontal lobe, and parietal lobe. In contrast, patients in the Epilepsiae dataset mostly have hippocampal sclerosis and temporal lobe epilepsy, and the brain regions of origin are mainly distributed in the limbic system, temporal lobe, and frontal lobe. The bar and error bar represent the mean and s.e.m., respectively. Note that the limbic system includes the hippocampus and the amygdala in this result.

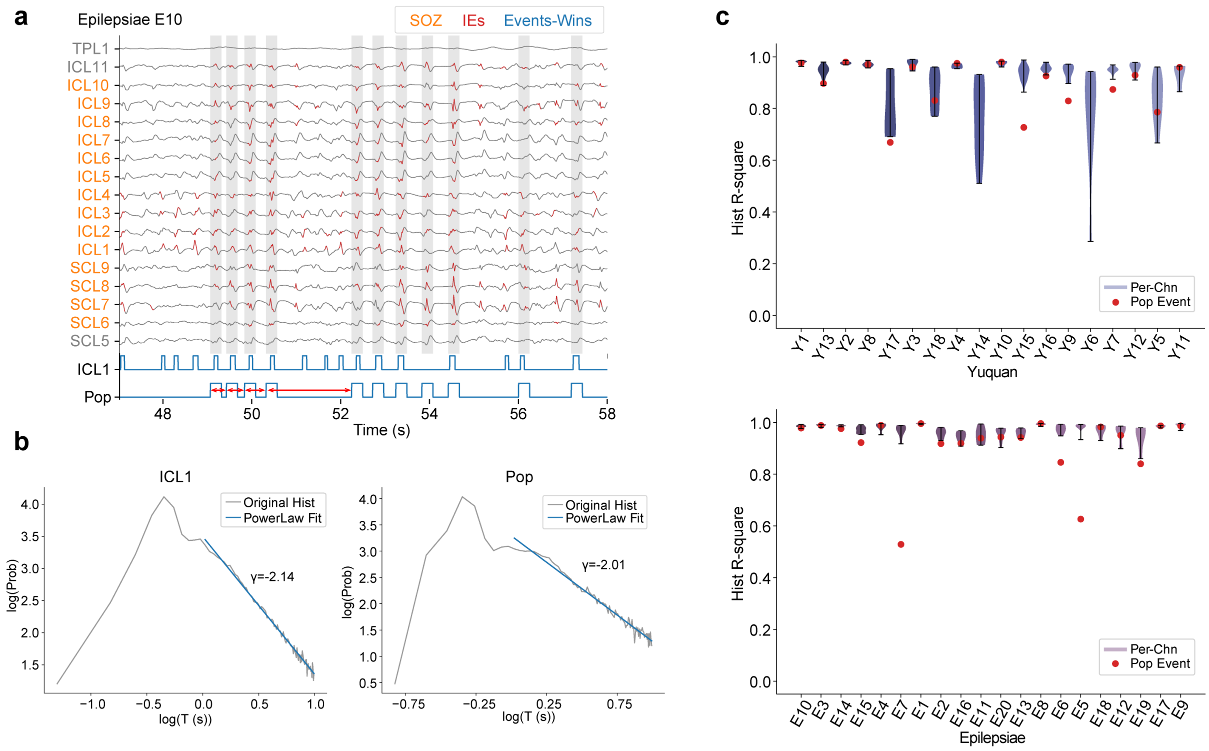


**Supplementary Fig. 7 | The time intervals between adjacent interictal events show a power-law distribution. a,** The red arrow lines in the time window signals at the bottom indicate the time intervals between adjacent interictal events considered in this analysis. **b,** For population activity or local activity at the ICL1 contact, a histogram represents the distribution of time intervals between adjacent events. Both the time intervals and the number of statistics are plotted logarithmically on the horizontal and vertical axes, respectively. The distribution and fitting results show that the time intervals follow a power-law distribution. **c,** The quality of the power-law fit to the distribution of time intervals (represented by the $R^2$ value of the fit). High $R^2$ values from the power-law fit indicate that the distributions of the inter-event intervals broadly exhibit power-law characteristics.

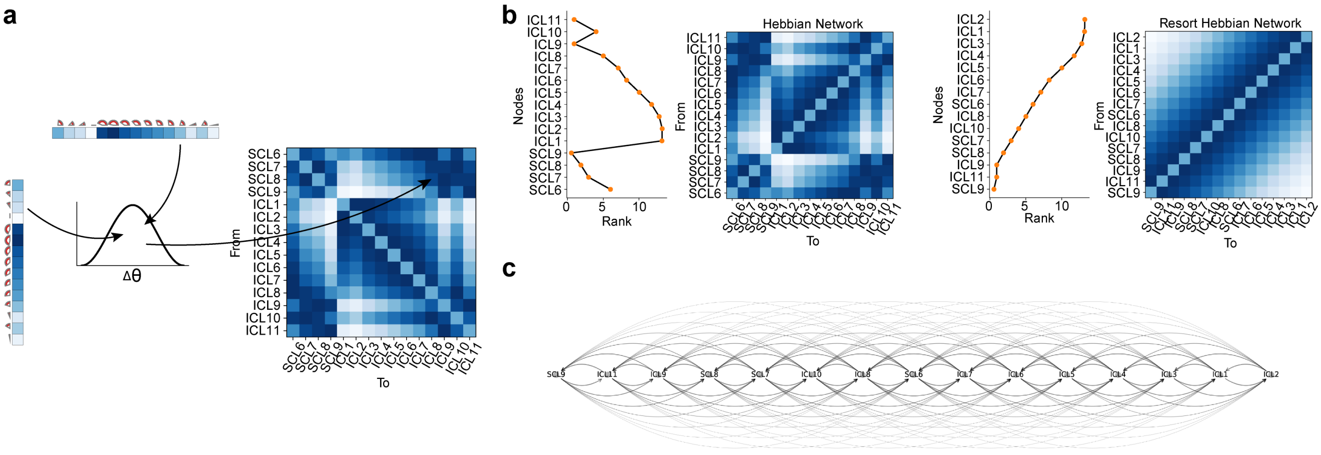


**Supplementary Fig. 8 | The interictal Hebb network structure has a linear recurrent structure related to the propagation pattern. a,** Based on Hebbian rule $A_{ij} = \cos(\Delta\theta_{ij})$, the phase patterns of the nodes are encoded into the interictal network structure. **b,** The interictal Hebb network structure is rearranged according to the order of propagation, with each set of figures showing the propagation pattern on the left and the corresponding network structure on the right. **c,** After rearrangement, the interictal network shows a linear recurrent structure.

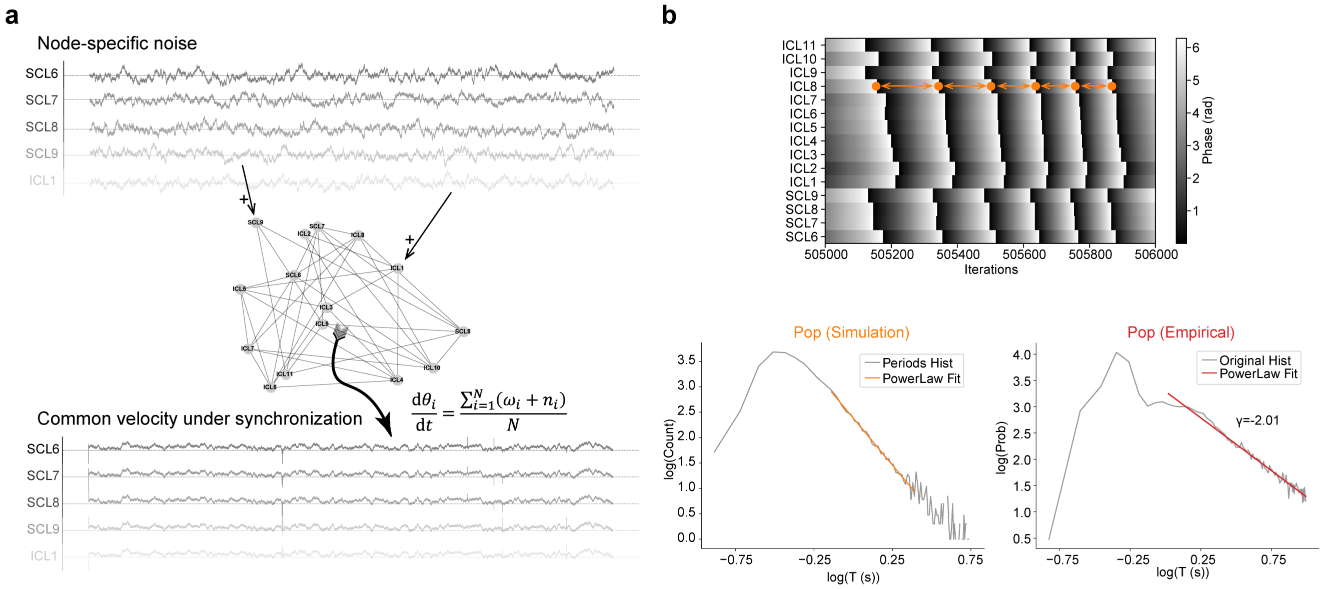


**Supplementary Fig. 9 | Implementing the Ornstein-Uhlenbeck Process (OUP) noise into the Kuramoto model can reproduce the power-law characteristics of the inter-event interval distribution in the data. a,** When independent OUP noise is applied to each node, the nodes exhibit a consistent velocity change trend under the synchronization effect of the Kuramoto model. **b,** Driven by the uniform collective velocity of the nodes in **a**, the distribution of the inter-event intervals (periods, shown as orange double-arrow lines) of network activity exhibits a power-law characteristic that is consistent with the data.

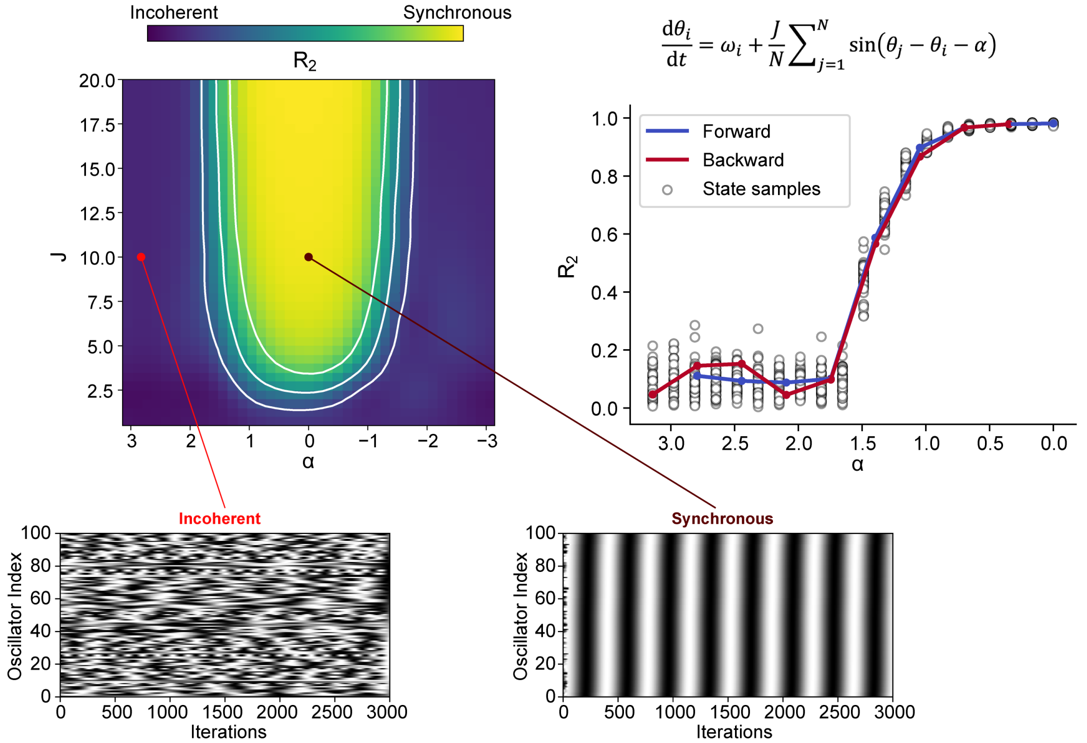


**Supplementary Fig. 10 | Dynamical analysis of an original Kuramoto model.** The original Kuramoto model is characterized by two key parameters, namely the phase delay $\alpha$ and the inter-node coupling strength $J$. By varying these two parameters, the system can exhibit different degrees of synchronization ($R_2$), including two main types: incoherent and synchronous. Unlike the adaptive Kuramoto model, the original Kuramoto model exhibits a continuous second-order phase transition, where the $R_2$ change curves for opposite directions overlap.

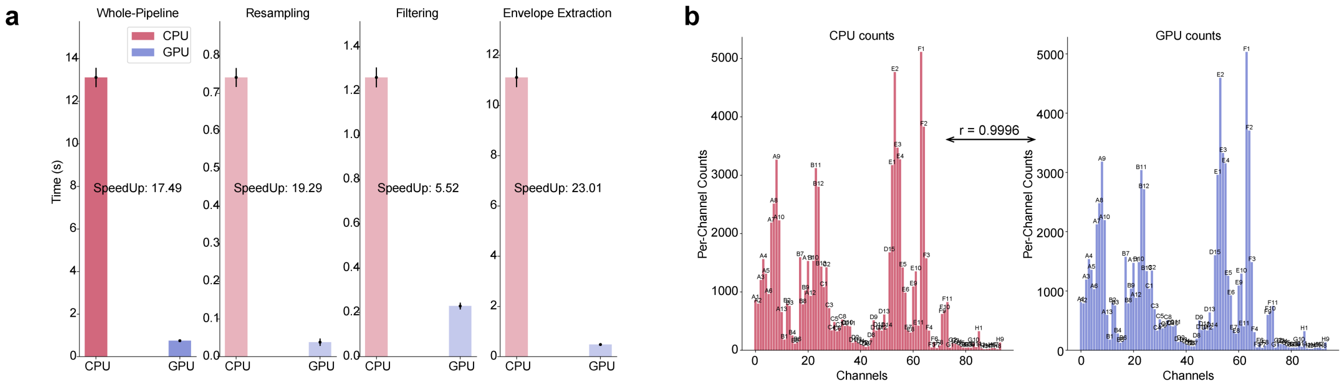


**Supplementary Fig. 11 | The acceleration effect and reliability of interictal event detection using GPU. a**, The acceleration effect of GPU compared to CPU at different stages of data processing. The comparison is based on 200-second segments in 2-hour intracranial EEG recording. The bar represents the mean, and the error bar represents the s.e.m. **b**, The consistency of interictal event detection between GPU and CPU on the same EEG data is high, with a Pearson correlation coefficient of 0.9996.

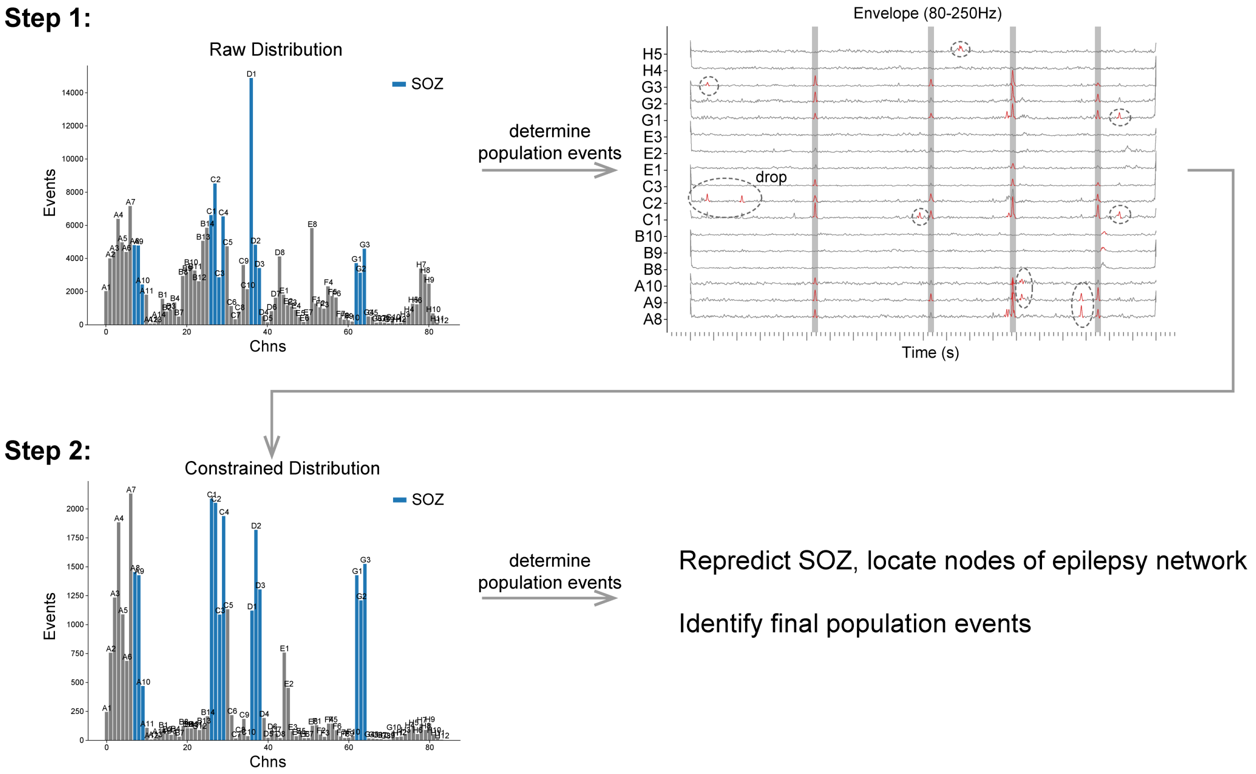


**Supplementary Fig. 12 | A two-stage optimization method based on synchronicity constraints of interictal events.** In the first step, raw distribution of interictal events are used for determining the population activity. In the second step, only interictal events participating in the population activity are considered, thereby eliminating the influence of physiological high-frequency or noise components. Based on the updated distribution of interictal events, further predictions regarding the SOZ or final population activity are made.

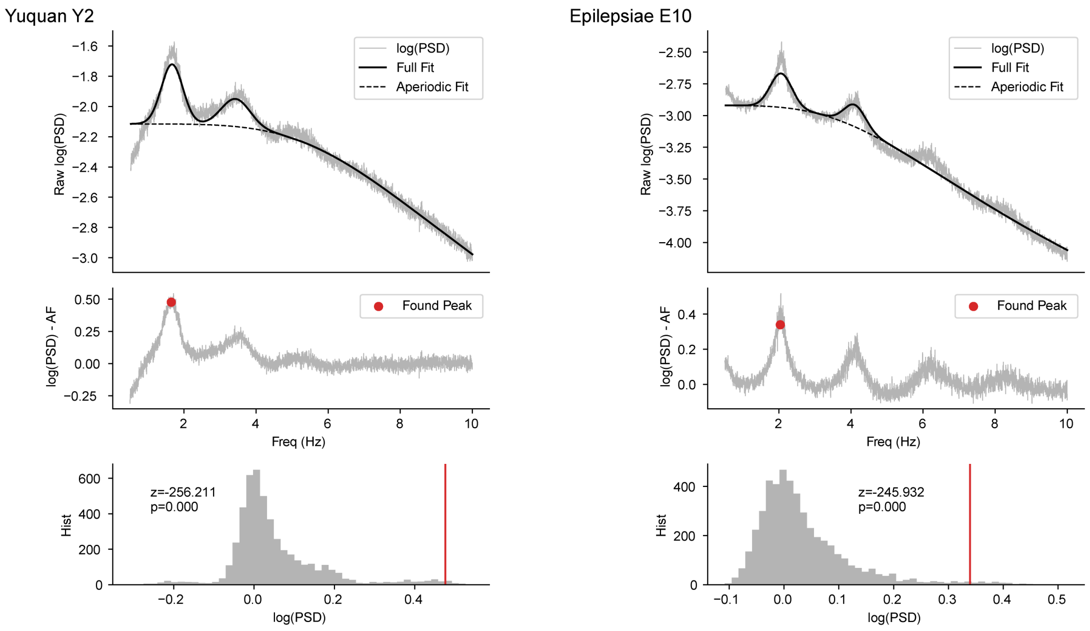


**Supplementary Fig. 13 | Significance test of periodicity of interictal events.** By conducting spectral fitting and decomposition of the occurrence signal of interictal events, both periodic and non-periodic components can be obtained. By removing the non-periodic components, we can then test the significance of the energy at the main periodic frequency positions obtained from the periodic fitting using t-test.

**Supplementary Table 1 | Yuquan patients information.**

| Patients | Gender | Age Range /year | Data Duration/hour | Fs/Hz | Contacts | Seizures | Etiology | Follow up |
|---|---|---|---|---|---|---|---|---|
| Y1 | F | 0-5 | 24 | 2000 | 102 | 3 | TSC | No seizures for 4 months after laser ablation |
| Y2 | M | 0-5 | 24 | 2000 | 148 | 6 | TSC | No seizures for 1 year after thermal coagulation |
| Y3 | M | 6-10 | 24 | 2000 | 140 | 3 | FCD | No seizures for 1 year after surgery |
| Y4 | F | 6-10 | 24 | 2000 | 158 | 2 | TSC | Laughing at 6-month follow-up after thermal coagulation |
| Y5 | F | 0-5 | 24 | 2000 | 102 | 2 | TSC | Still having seizures after laser ablation |
| Y6 | M | 0-5 | 24 | 2000 | 206 | 17 | TSC | Relapse after 8 months of thermocoagulation, but in reduced numbers |
| Y7 | F | 0-5 | 22 | 2000 | 172 | 1 | FCD | No seizures for 1 and a half years after surgery |
| Y8 | F | 6-10 | 26 | 2000 | 142 | 3 | TSC | No seizures for 7 months after surgery |
| Y9 | F | 6-10 | 24 | 2000 | 114 | 3 | FCD | No seizures for 8 months after thermal coagulation |
| Y10 | F | 16-20 | 26 | 2000 | 144 | 4 | ganglioglioma | No seizures after surgery, no follow-up |
| Y11 | M | 0-5 | 24 | 2000 | 118 | 2 | TSC | Improvement after laser ablation |
| Y12 | M | 11-15 | 24 | 2000 | 112 | 1 | TSC | No seizures for 8 months after thermal coagulation |
| Y13 | M | 21-25 | 26 | 2000 | 148 | 1 | FCD | No seizures for 1 year after thermal coagulation |
| Y14 | M | 6-10 | 24 | 2000 | 94 | 1 | FCD | No seizures for 1 and a half years after surgery |
| Y15 | M | 16-20 | 32 | 2000 | 142 | 8 | FCD | No seizures for 1 year after thermal coagulation |
| Y16 | F | 6-10 | 26 | 2000 | 130 | 3 | TSC | Relapse after 7 months of thermal coagulation |
| Y17 | F | 0-5 | 26 | 2000 | 104 | 8 | FCD | No seizures for 8 months after thermal coagulation |
| Y18 | M | 0-5 | 24 | 2000 | 128 | 8 | TSC | No seizures for 5 months after thermal coagulation |

**Supplementary Table 2 | Epilepsiae patients information.**

| Patients | Gender | Age Range /year | Data Duration/hour | Fs/Hz | Contacts | Seizures | Etiology | Follow up |
|---|---|---|---|---|---|---|---|---|
| E1 | F | 31-35 | 162.6 | 1024 | 100 | 9 | malformation, hippocampal sclerosis | Ia (3m) |
| E2 | F | 46-50 | 244.7 | 1024 | 117 | 94 | malformation | Ia (3m)<br>IIb (11m) |
| E3 | F | 11-15 | 219.1 | 1024 | 121 | 16 | cryptogenic | Ia (3m) |
| E4 | M | 36-40 | 110.6 | 1024 | 107 | 30 | cryptogenic | Ia (3-24m) |
| E5 | M | 16-20 | 245.2 | 1024 | 122 | 13 | malformation | Ia (3-24m) |
| E6 | F | 31-35 | 151.6 | 1024 | 125 | 9 | cryptogenic | IVb (3-6m) |
| E7 | M | 21-25 | 170.6 | 1024 | 95 | 22 | genetic risk | Ia (3-12m) |
| E8 | F | 46-50 | 224.0 | 1024 | 96 | 20 | hippocampal sclerosis | IIIa (3-12m) |
| E9 | F | 36-40 | 260.1 | 512 | 58 | 7 | hippocampal sclerosis | IIb (3m)<br>Ia (48m) |
| E10 | M | 21-25 | 113.2 | 1024 | 116 | 26 | vascular hypoxia | Not offered |
| E11 | M | 21-25 | 424.1 | 1024 | 109 | 52 | malformation | Ib (3-24m) |
| E12 | M | 41-45 | 252.4 | 1024 | 63 | 7 | malformation | Ia (3m) |
| E13 | F | 21-25 | 63 | 1024 | 65 | 23 | malformation, tumor | Ia (3m)<br>IIa (36m) |
| E14 | M | 16-20 | 142.0 | 1024 | 109 | 31 | malformation, hippocampal sclerosis | Ia (3m-24m) |
| E15 | F | 45-50 | 65 | 1024 | 119 | 15 | hippocampal sclerosis | IIb (3-6m) |
| E16 | F | 51-55 | 130 | 512 | 63 | 6 | hippocampal sclerosis | IVb (3m)<br>Ia (60m) |
| E17 | F | 11-15 | 155.0 | 1024 | 87 | 14 | malformation, hippocampal sclerosis | Ia (3m) |
| E18 | F | 26-30 | 183.1 | 1024 | 122 | 9 | malformation, hippocampal sclerosis | Ia (3m) |
| E19 | F | 26-30 | 248.7 | 1024 | 71 | 9 | hippocampal sclerosis | Ib (3m)<br>IIIa (24m) |
| E20 | F | 61-65 | 118.9 | 1024 | 82 | 21 | tumor | Not offered |
